# Performance of ICD10 Code-Based Dementia Case Definition in the Health and Retirement Study

**DOI:** 10.64898/2026.01.12.26343791

**Authors:** Kan Z. Gianattasio, Michael Steffan, Ben Reist, Melinda C. Power, David Rein

## Abstract

**INTRODUCTION:** The Dementia DataHub (DDH) reports U.S. dementia prevalence and incidence from Medicare data. Variation in sensitivity, specificity, and accuracy of diagnoses related to geography and participant characteristics complicates interpretation of these data.

**METHODS:** We evaluated performance of DDH-defined Medicare claims diagnoses against linked HRS survey-based dementia classifications.

**RESULTS:** DDH’s likely-or-higher dementia definition achieved 50% sensitivity, 97% specificity, and 91% accuracy relative to the HRS classification. Sensitivity was higher in urban settings, but patterns varied across census divisions. Respondents with dementia missed in claims were younger and healthier than those correctly identified.

**DISCUSSION:** Medicare claims reflecting diagnoses of dementia provide valuable information about who may be receiving dementia treatment and where; however, they often miss cases in ways that differ by geography and patient characteristics. Variation in diagnosis in Medicare claims relative to HRS survey-based dementia classification can be used to improve the value of Medicare diagnoses for surveillance purposes.

## BACKGROUND

Alzheimer’s disease and Alzheimer’s disease related dementias (AD/ADRD) impose substantial health, caregiving, and financial burdens upon older adults in the United States.^1,2^ Until recently, no national surveillance system tracked the diagnosis of AD/ADRD in the U.S., limiting our ability to understand the distribution of AD/ADRD burden and plan care accordingly.

To address this critical surveillance gap, the *Dementia DataHub* (DDH), an online data portal with public use files accessible at https://www.dementiadatahub.org/, disseminates data on the prevalence and incidence of diagnosed dementia at the state and county levels.^3^ DDH estimates are derived by applying NORC’s researcher-consensus and data-driven ICD-10 and National Drug Code (i.e. code-based) AD/ADRD case definition to Medicare fee-for-service (FFS) claims and Medicare Advantage (MA) encounter data.^4^ Medicare claims are a strong source of data for measuring AD/ADRD because Medicare covers nearly all older adults in the U.S. However, claims may not accurately capture all persons with AD/ADRD due to underdiagnosis of dementia in clinical settings, overdiagnosis of persons exhibiting mild cognitive difficulties, and variations in diagnosis and coding practices across region and clinical settings.^5^ While Medicare claims provide a highly accurate accounting of individuals who have received specific AD/ADRD diagnosis codes and dementia targeting drugs in their billing records, the extent to which this diagnostic signal reflects the underlying prevalence of all AD/ADRD in a geography is unknown. This complicates the use of claims data for surveillance, because geographic and/or characteristic-related variations in AD/ADRD diagnoses do not necessarily reflect true variations in AD/ADRD prevalence or incidence.

Comparing Medicare diagnostic information to other independently collected measures of cognitive function in a geographically distributed and U.S.-representative sample can uncover differences in the likelihood of receiving a correct diagnosis; this can subsequently reveal differences in diagnosis rates by geography and personal characteristics, including demography, insurance category, health status, and cognitive capacity. In this paper, we used linked Health and Retirement Study (HRS)-Medicare data to examine performance of the DDH’s code-based case definitions as compared to dementia status determined probabilistically using previously published HRS algorithms.^6^ Findings from this study can be used in the future to adjust estimates observed in Medicare to improve their accuracy in capturing variation in diagnosed and undiagnosed dementia prevalence and incidence by geography and individual characteristics.

## METHODS

### Data Sources

The HRS, which began in 1992, is a nationally representative longitudinal survey of older adults (aged 50+); it has maintained a steady-state sample by replenishing participants every six years since 2004. Interviews are conducted every two years, collecting demographic, health, and socioeconomic data from participants.^7^ The HRS follows individuals through death, and allows proxies to complete the interview for participants who are unable or unwilling, reaching approximately 19,000 respondents at each wave.^8,9^ We used data from the 2018 wave for this study.

Medicare fee-for-service (FFS) claims and Medicare Advantage (MA) encounter data contain information on service dates, as well as primary and secondary ICD-10 diagnosis codes. We used the Medicare master beneficiary summary files (MBSF); FFS inpatient, outpatient, carrier, hospice, skilled nursing facility (SNF), home health claims data; MA inpatient, outpatient, carrier, home health, SNF encounter data; and part D claims data between 2016 and 2018 in this study.

### Sample

We excluded HRS participants aged under 65 at the time of their 2018 wave HRS interview (n=8,129), those without Medicare enrollment or who did not have 2018 Medicare linkage (n=1,032), and those with missing race/ethnicity (n=4) for a total of 7,981 participants.

### HRS-identified dementia

The HRS does not conduct formal dementia assessments, but HRS interviews include cognitive testing data, which, along with other health and sociodemographic information, has been used to predict dementia status as compared to gold standard measures. Researchers have developed a number of such algorithms by leveraging dementia status ascertained through formal assessments for the 856 participants sampled in the Aging, Demographics, and Memory Study (ADAMS) sub-study.^6,10^ In this study, we applied three algorithms developed with the explicit intent to minimize differences in model performance across race/ethnicity groups (Expert, LASSO, and Modified-Hurd); each algorithm computes a dementia probability, and applies race/ethnicity-specific score cut-offs to designate dementia.^6^ Due to sporadic missingness of variables specific to each HRS algorithm, the analytic sample size varied: 7,901 using the Expert Model, 7,518 using the LASSO model, and 7,909 using the Modified-Hurd Model.

A number of survey items, including 9 self-respondent cognition assessment items (backwards counting, year, month, day of week, date recall, cactus, scissor, president, and vice president naming) used as inputs in the dementia prediction algorithms were missing for a random subset of self-respondent, English-speaking, community-dwelling respondents assigned to web-mode interviews^11,12^ We used imputed values published by the University of Michigan for missing backwards counting data,^11^ and applied multiple imputation chained equations (MICE) to produce 10 sets of imputations for date recall and item/president/VP naming (details in **Appendix 1**).

### Medicare-identified dementia

We identified all 2016-2018 Medicare FFS claims and MA encounters with at least one ICD-10 code from the code-based dementia case definition, and all 2016-2018 part D drug claims with a prescription for an AD/ADRD-targeting drug.^4^ Respondents with at least two claims/encounters originating on separate dates with a highly likely/likely code were categorized as having highly likely dementia; those with one corresponding claim/encounter were categorized as having likely dementia; and those with at least one claim/encounter with a possible code or at least one AD/ADRD-targeting drug prescription were categorized as having possible dementia. We consider two definitions in this paper: the likely-or-higher definition combining likely and highly likely dementia categories, and the possible-or-higher definition combining all three categories.

### Statistical analysis

We compute sensitivity, specificity, and accuracy of DDH-defined dementia from Medicare claims against algorithmic HRS dementia. We also computed the distribution of true positive (TP; positive HRS and DDH), false negative (FN; positive HRS and negative DDH), true negative (TN; negative HRS and DDH), and false positive (FP; negative HRS and positive DDH) classifications. We stratified our sample by FFS- vs. MA-enrolled participants, by rurality based on 2023 Beale rural-urban code, and by U.S. census divisions (variable specifications in **Appendix 2**) to evaluate differences in performance across insurance and geography. To understand differences in diagnostic accuracy across participant characteristics, we compared the four classification groups (TP, FN, TN, FP) in terms of their predicted HRS dementia probabilities – considered as a composite measure of dementia “risk” –and their HRS-measured demographic, insurance, physical health, and cognitive characteristic distributions. We focused on differences between the true positive vs. false negative groups to evaluate drivers of underdiagnosis, and between the true negative vs. false positive groups to evaluate drivers of overdiagnosis.

Finally, we investigated differences in how specific ICD-10 codes perform in identifying dementia. We first counted the number of participants who were uniquely diagnosed by each ICD-10 code. Because there were few participants identified uniquely by any single code, we grouped codes with shared “roots” (e.g. all codes beginning with F03, which fall under “Unspecified dementia”) within the high likely/likely and within the possible categories. To further ensure adequate sample sizes, we combined across roots, whereby uniqueness was then defined at the group level (e.g. combining F01 and F02 codes allowed participants to have both F01 and F02 codes in their claims). We then computed, for each unique code group, the proportion of true positives vs. false positives.

We considered the Expert Model as the primary outcome, and the LASSO and Modified-Hurd Models as sensitivity analyses. Unless otherwise specified, we also considered the code-based likely-or-higher definition as the primary outcome, and the possible-or-higher definition as sensitivity analyses. As an additional sensitivity check, we re-estimated analyses excluding web-respondents for whom we had to impute values for date, object, and president/VP-naming cognitive assessments.

All analyses use complex variance estimation to account for the HRS survey design. For all outcomes, we report means across the 10 imputations for missing web-respondent cognitive data and standard errors that account for both within and between-imputation variance.

## RESULTS

The 2018 HRS analytic sample with linked CMS and non-missing Expert Model dementia classification contained 7,901 respondents, representing 45.9 million individuals, with the majority being female (55.5%), non-Hispanic White (80.5%), and aged under 75 (54.6%). The sample was generally in good physical health, with just 27.3% reporting their health to be poor or fair, and fewer than 20% reporting at least 1 activity of daily living (ADL) or instrumental activity of daily living (IADL) limitation; only 1.7% died during the year. Most of the sample had FFS coverage (59.0%) and were non-dual eligible (87.5%). Geographically, the majority resided in an urban setting (72.4%), and representation by census division ranged from 4.5% in New England to 23.4% in the South Atlantic. The prevalence of dementia was estimated at 11.6% to 12.0% across the three HRS probabilistic algorithms (descriptive statistics and confidence intervals in **Appendix 3**).

Overall, the DDH likely-or-higher definition achieved a sensitivity of 50%, specificity of 97%, and overall accuracy of 91% (**Table 1**). Of the HRS sample, the majority (85.4%) were correctly identified as having no dementia (i.e. true negative), 6.0% were true positive, 6.0% were false negative, and 2.7% were false positive. Across insurance programs, codes correctly identified a higher proportion of HRS-positive dementia in FFS beneficiaries than in MA beneficiaries (52% vs. 46% sensitivity, p = 0.01), with no loss in specificity (97%). Code-based dementia did not differ significantly in sensitivity by rurality (51% in rural vs. 48% in urban settings, p = 0.31) but was marginally more specific in rural than urban settings (98% vs. 97%, p = 0.03). Including possible dementia yielded an overall sensitivity of 59%, specificity of 93%, and accuracy of 89%. No significant differences in performance were observed by insurance type or rurality.

**Table 1.** Performance of DDH Dementia Case Definition Against the HRS Expert Model.

| Sample | Performance Metrics (95% CI) <sup>a</sup> |  |  | Classification Distribution (95% CI) <sup>a</sup> |  |  |  |
| --- | --- | --- | --- | --- | --- | --- | --- |
|  | Sensitivity | Specificity | Accuracy | True positive | False negative | True negative | False positive |
| *Likely-or-higher dementia* |  |  |  |  |  |  |  |
| Overall | 50 (46 - 53) | 97 (96 - 97) | 91 (91 - 92) | 6 (5.3 - 6.6) | 6 (5.4 - 6.6) | 85.4 (84.3 - 86.4) | 2.7 (2.3 - 3.1) |
| Insurance type |  |  |  |  |  |  |  |
| FFS | 52 (48 - 57) | 97 (96 - 98) | 92 (91 - 93) | 6.2 (5.3 - 7) | 5.6 (4.8 - 6.4) | 85.7 (84.2 - 87.2) | 2.6 (2.1 - 3.2) |
| MA | 46 (42 - 50) | 97 (96 - 98) | 91 (89 - 92) | 5.7 (4.9 - 6.4) | 6.6 (5.7 - 7.5) | 84.9 (83.5 - 86.3) | 2.8 (2.1 - 3.5) |
| p-value <sup>b</sup> | 0.01 (0.01 - 0.02) | 0.67 (0.57 - 0.68) | 0.12 (0.11 - 0.19) | 0.08 (0.06 - 0.15) |  |  |  |
| Setting |  |  |  |  |  |  |  |
| Rural | 48 (43 - 53) | 98 (97 - 98) | 91 (89 - 92) | 6.9 (5.8 - 8.1) | 7.6 (5.9 - 9.2) | 83.6 (81.2 - 86) | 1.9 (1.3 - 2.5) |
| Urban | 51 (47 - 55) | 97 (96 - 97) | 92 (91 - 92) | 5.6 (4.8 - 6.4) | 5.4 (4.8 - 6) | 86 (84.9 - 87.2) | 3.0 (2.4 - 3.5) |
| p-value <sup>b</sup> | 0.31 (0.25 - 0.39) | 0.03 (0.03 - 0.04) | 0.23 (0.19 - 0.28) | 0.02 (0.01 - 0.02) |  |  |  |
| *Possible-or-higher dementia* |  |  |  |  |  |  |  |
| Overall | 58 (55 - 62) | 93 (92 - 93) | 89 (88 - 90) | 7 (6.3 - 7.7) | 5 (4.3 - 5.6) | 81.6 (80.3 - 82.8) | 6.5 (5.8 - 7.2) |
| Insurance type |  |  |  |  |  |  |  |
| FFS | 60 (56 - 64) | 93 (92 - 94) | 89 (88 - 90) | 7 (6.2 - 7.9) | 4.7 (3.9 - 5.4) | 82 (80.3 - 83.6) | 6.3 (5.5 - 7.1) |
| MA | 56 (52 - 61) | 92 (91 - 94) | 88 (87 - 89) | 6.9 (6.1 - 7.8) | 5.4 (4.5 - 6.2) | 81 (79.3 - 82.7) | 6.7 (5.6 - 7.8) |
| p-value <sup>b</sup> | 0.11 (0.09 - 0.18) | 0.51 (0.43 - 0.54) | 0.22 (0.18 - 0.26) | 0.42 (0.34 - 0.53) |  |  |  |
| Setting |  |  |  |  |  |  |  |
| Rural | 55 (48 - 62) | 93 (92 - 95) | 88 (85 - 90) | 8 (6.5 - 9.5) | 6.5 (4.8 - 8.2) | 79.7 (76.7 - 82.8) | 5.8 (4.4 - 7.2) |
| Urban | 60 (56 - 64) | 92 (92 - 93) | 89 (88 - 90) | 6.6 (5.8 - 7.4) | 4.4 (3.9 - 4.9) | 82.3 (81.1 - 83.5) | 6.7 (6 - 7.5) |
| p-value <sup>b</sup> | 0.22 (0.19 - 0.26) | 0.38 (0.36 - 0.41) | 0.35 (0.32 - 0.39) | 0.03 (0.02 - 0.04) |  |  |  |

Abbreviations: CI = confidence interval; DDH = Dementia Datahub; FFS = fee-for-service Medicare; HRS = Health and Retirement Study; MA = Medicare Advantage.
<sup>a</sup> Values in paratheses represent 95% CI except for p-values, for which values in parentheses represent minimum and maximum of estimates obtained across the 10 imputed datasets.
<sup>b</sup> Reported p-values represent the median (minimum - maximum) of estimates obtained across the 10 imputed datasets. P-values indicate significance in differences between subgroups (FFS vs. MA; Rural vs. Urban).
Notes: A subset of self-report cognitive data were missing for wave 2018 web-respondents (11.2% of the sample); we used multiple imputation chained equations (MICE) to generate 10 sets of imputations for these missing data. Point estimates were averaged across the ten datasets, and confidence intervals incorporate both within- and between-imputation variability. PPV/NPV are omitted because they vary with subgroup prevalence and are not comparable across subgroup populations.
All analyses use complex variance estimation to account for the HRS survey design.

Geographically, the sensitivity of code-based dementia varied substantially across census divisions (p = 0.06), at approximately 20 percentage points higher in the East South Central (60%) and Mid Atlantic (56%) divisions than in the Pacific (36%) and West North Central (43%) regions; specificity varied to a lesser degree, ranging from 94% to 98% (p = 0.01) (**Figure 1**; **Appendix 4**). Performance further varied by rural-urban setting and census divisions: most notably, code-based dementia was both more sensitive and specific in urban settings in New England; less sensitive and less specific in urban settings in Mid Atlantic and South Atlantic; and less sensitive but marginally more specific in urban settings in the Pacific. Accuracy was highest in the East South Central (94%) and Mid Atlantic (93%) divisions, and was higher in urban settings in five of the nine divisions, with the largest difference occurring in the West South Central division (7 percentage points).

**Figure 1.**
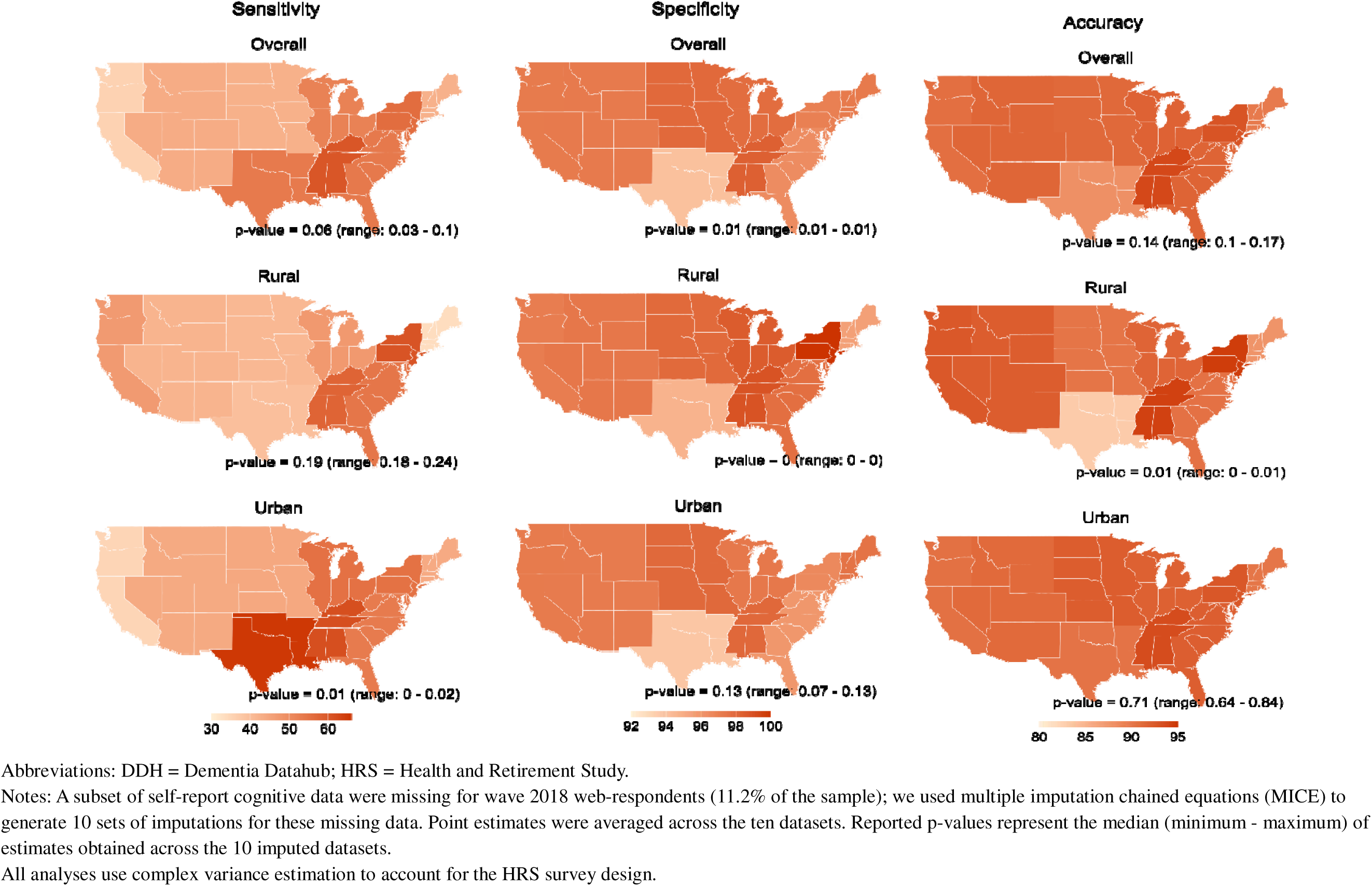
Performance of DDH likely-or-higher dementia against HRS Expert Model dementia classification by Census division and rural/urban setting.

Among only the DDH highly likely dementia category, the mean probability of dementia estimated using the HRS Expert Model was 0.82 for true positives, compared to 0.62 for false negatives, with some overlap in inter-quartile range (IQR) (**Figure 2**; **Appendix 5**). Conversely, the mean probability of dementia was 0.02 for true negatives, and 0.08 for false positives. Estimated dementia probabilities across the true positive, false negative, and false positive groups shifted slightly lower when considering the likely-or-higher definition, and more so when considering the possible-or-higher dementia category. IQR became wider among true positives and narrower among false positives moving from highly likely to possible-or-higher claims-based dementia.

**Figure 2.**
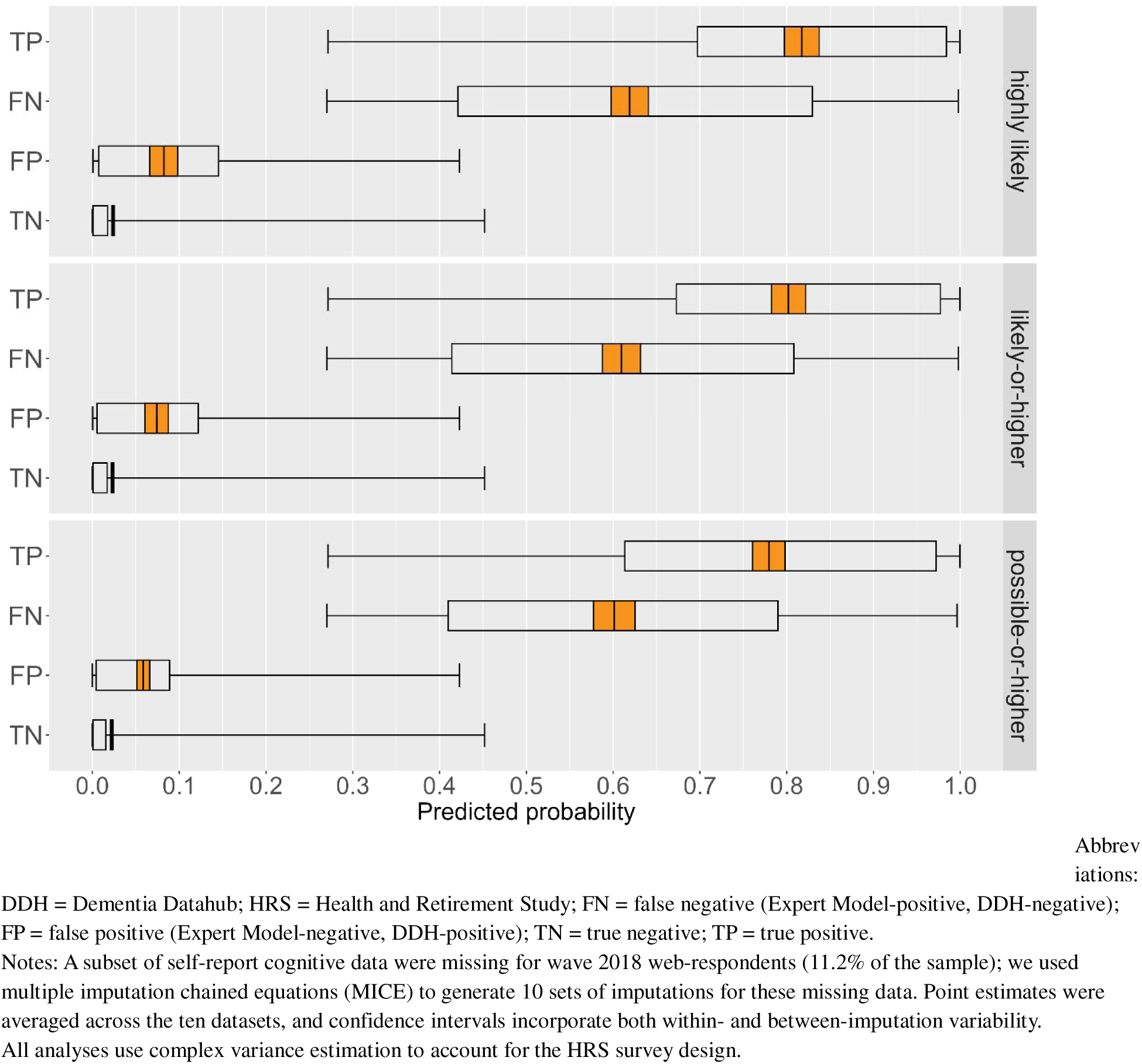
Dementia probabilities estimated using the HRS Expert Model for participants classified as having DDH highly likely, likely-or-higher, and possible-or-higher dementia.

Among those with and those without HRS-based dementia, we found lower cognitive ability, as measured by both self- and proxy-respondent instruments, in individuals with DDH classified dementia, indicating lower risk of under-diagnosis but higher risk of over-diagnosis (**Table 2**). Similarly, individuals with at least an ADL limitation or an IADL limitation, those who died in the reference year and those who were dually-eligible for Medicare and Medicaid had lower risk of under-diagnosis if they have HRS-based dementia (i.e. over-represented in the true positive relative to the false negative group), but higher risk of over-diagnosis if they do not have HRS-based dementia (i.e. over-represented in the false positive relative to the true negative group). Conversely, women and individuals with a diagnosis of high blood pressure or diabetes had higher HRS-based dementia prevalence than their counterparts but were less likely to have evidence of claims-based dementia conditional on having HRS-based dementia, indicating higher risk of under-diagnosis.

**Table 2.**
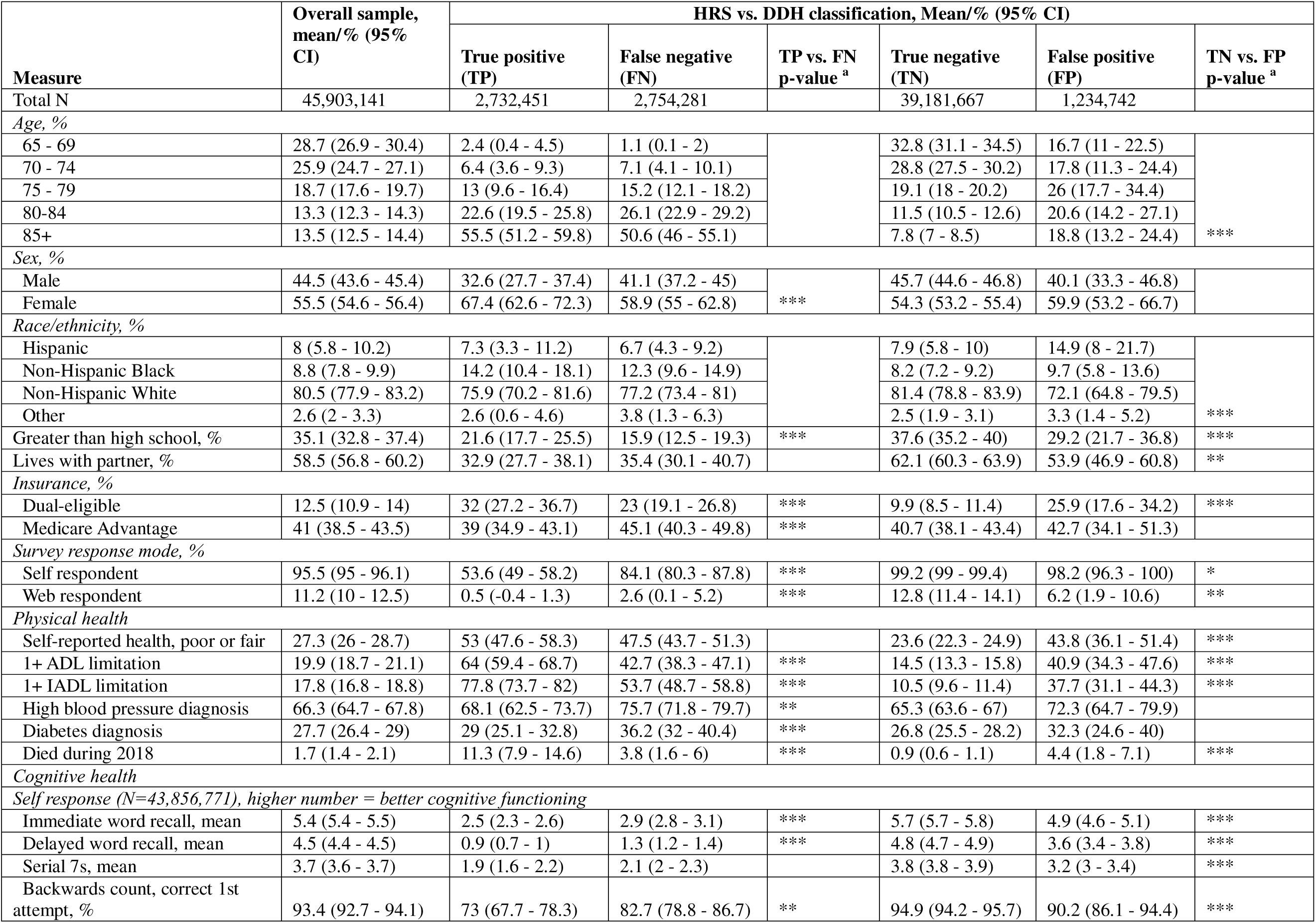
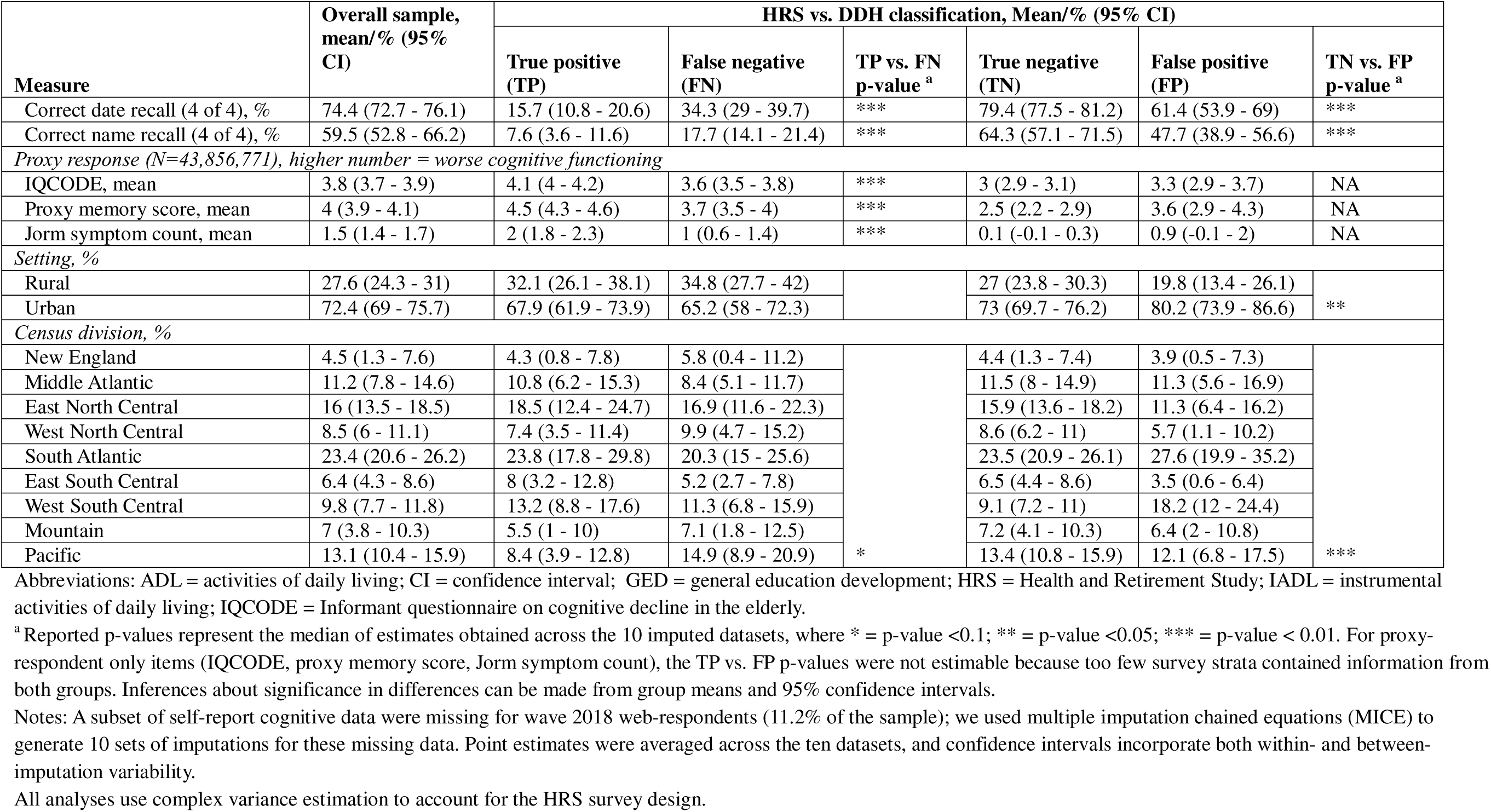
Sample Descriptive Statistics, Overall and by HRS Expert Model and DDH likely-or-higher dementia classifications.

Among those without HRS-based dementia, greater age, having poor or fair self-reported health, living in an urban setting, and Hispanic ethnicity were associated with greater risk of over-diagnosis, while non-Hispanic white race was associated with lower risk of over-diagnosis. There appeared to be no differences in misdiagnosis among those with HRS-based dementia across race/ethnicity groups. Finally, having a greater than high school education was associated with both fewer false negatives and fewer false positives.

We found that those with likely-or-higher dementia were most likely to be uniquely diagnosed with F03 codes (unspecified dementia), which alone had a true positive rate of 59% (**Table 3**). Participants were unlikely to be uniquely diagnosed with only F01 (vascular dementia), F02 (Dementia in other diseases classified elsewhere), or G30 (Alzheimer’s dementia) codes; in combination, the three codes had a true positive rate of 53%. All “possible” category dementia codes had substantially lower true positive rates, ranging 17% for G31.9 (Degenerative disease of nervous system, unspecified) to 31% for R54 (Age-related physical debility); excluding G31.9, G31.84 (mild cognitive impairment), and R54, the other possible codes combined had a true positive rate of just 14%.

**Table 3.**
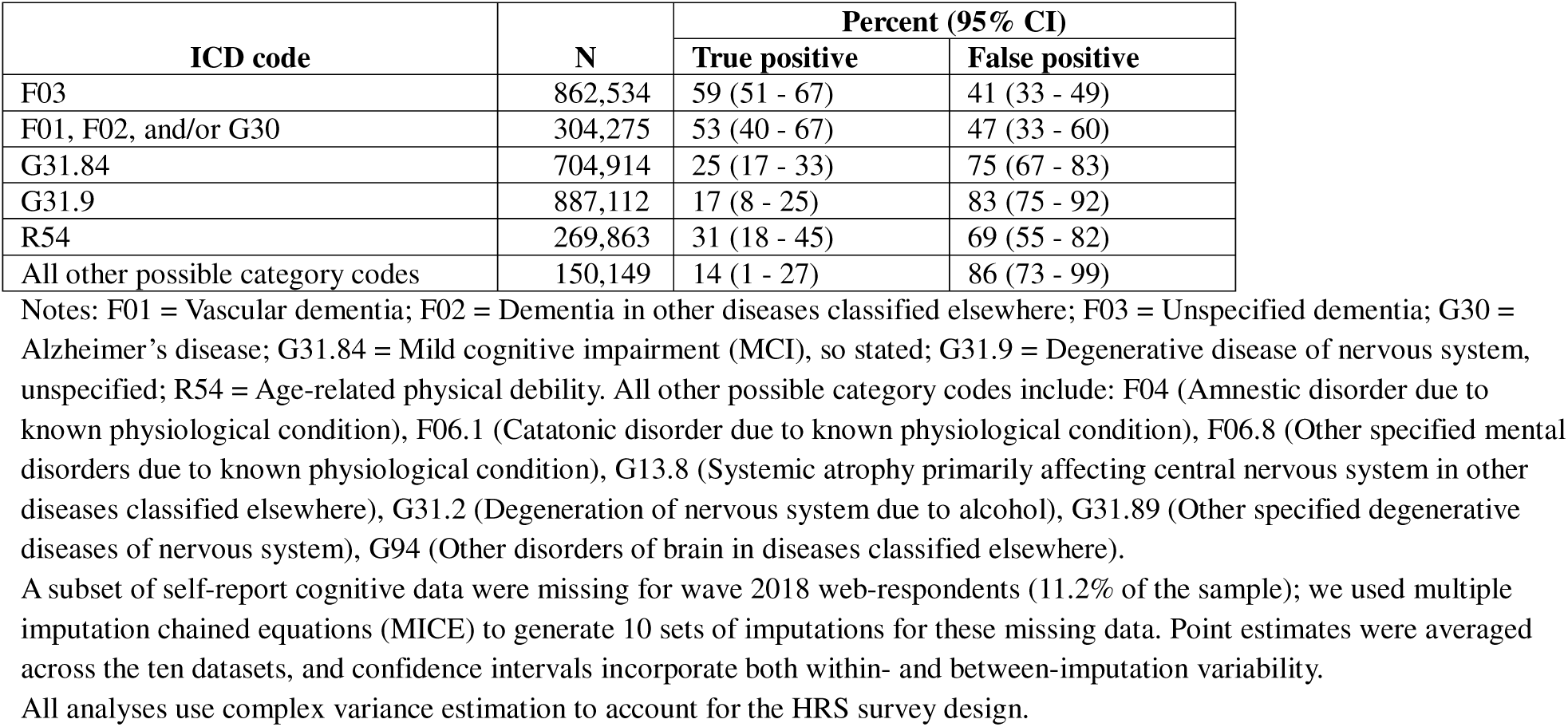
Performance of specific ICD-10-CM Codes Against HRS Expert Model Dementia.

| ICD code | N | Percent (95% CI) |  |
| --- | --- | --- | --- |
|  |  | True positive | False positive |
| F03 | 862,534 | 59 (51 - 67) | 41 (33 - 49) |
| F01, F02, and/or G30 | 304,275 | 53 (40 - 67) | 47 (33 - 60) |
| G31.84 | 704,914 | 25 (17 - 33) | 75 (67 - 83) |
| G31.9 | 887,112 | 17 (8 - 25) | 83 (75 - 92) |
| R54 | 269,863 | 31 (18 - 45) | 69 (55 - 82) |
| All other possible category codes | 150,149 | 14 (1 - 27) | 86 (73 - 99) |

Notes: F01 = Vascular dementia; F02 = Dementia in other diseases classified elsewhere; F03 = Unspecified dementia; G30 = Alzheimer's disease; G31.84 = Mild cognitive impairment (MCI), so stated; G31.9 = Degenerative disease of nervous system, unspecified; R54 = Age-related physical debility. All other possible category codes include: F04 (Amnestic disorder due to known physiological condition), F06.1 (Catatonic disorder due to known physiological condition), F06.8 (Other specified mental disorders due to known physiological condition), G13.8 (Systemic atrophy primarily affecting central nervous system in other diseases classified elsewhere), G31.2 (Degeneration of nervous system due to alcohol), G31.89 (Other specified degenerative diseases of nervous system), G94 (Other disorders of brain in diseases classified elsewhere).
A subset of self-report cognitive data were missing for wave 2018 web-respondents (11.2% of the sample); we used multiple imputation chained equations (MICE) to generate 10 sets of imputations for these missing data. Point estimates were averaged across the ten datasets, and confidence intervals incorporate both within- and between-imputation variability.
All analyses use complex variance estimation to account for the HRS survey design.

Relative performance of the Medicare possible-or-higher dementia definition by census divisions and rural/urban setting differed slightly from that of the likely-or-higher dementia definition (**Appendix 6**). All other sensitivity analyses were qualitatively consistent with primary analyses (**Appendix 7**: Medicare likely-or-higher dementia performance evaluated against other HRS models and excluding web respondents; **Appendices 8 – 10:** Medicare likely-or-higher dementia performance by census division and rural/urban settings; **Appendices 11 – 13:** Probability of dementia estimated using other HRS models and excluding web respondents for participants classified as having Medicare highly likely, likely-or-higher, and possible-or-higher dementia; **Appendix 14**: Descriptive characteristics by HRS Expert Model and Medicare possible-or-higher dementia classifications; **Appendix 15**: Performance of specific ICD-10-CM codes).

## DISCUSSION

We tested the performance of DDH code-based dementia case definitions in Medicare claims and encounter data against algorithmically identified dementia status in the linked 2018 HRS data. Sensitivity was 50% for likely-or-higher dementia and 59% for possible-or-higher dementia, while specificity exceeded 90% in both definitions. These metrics indicate that a substantial proportion of dementia is undiagnosed in clinical settings, underscoring the difficulty of diagnosing dementia in a timely manner.^13,14^ We also found a high degree of geographic variation in claims sensitivity, which correspond with the variation in Medicare estimates of dementia prevalence published on *Dementia DataHub.*^3^ Therefore, observed variations in diagnosed Medicare prevalence are likely driven in part by differences in diagnosis and/or documentation in claims and encounter records.

As expected, individuals with a higher algorithmically predicted probability (or composite risk) of dementia in HRS, as well as those who had poorer cognitive and physical health had lower risk of under-diagnosis but higher risk of over-diagnosis. We also found older age to be associated with greater risk of over-diagnosis, which is consistent with a prior study of all-cause dementia in the Washington Heights-Inwood Columbia Aging Project study, which found older age to be associated with a false positive diagnosis.^15^ With regard to educational attainment, we found the DDH case definition to be more accurate among individuals with a greater than high school education – both in those with dementia (fewer false negatives) and in those without (fewer false positives) dementia, consistent with previous studies of code-based definitions.^16,17^ We hypothesize that higher educational attainment improves ability to recognize and communicate symptoms to facilitate timely diagnoses in true dementia cases, while at the same time providing greater cognitive reserve, reducing the likelihood of being incorrectly diagnosed with AD/ADRD among those without dementia. Finally, contrary to other studies, including one that similarly uses linked HRS-Medicare data,^16,18–21^ we did not find disproportionate representation of non-Hispanic blacks or Hispanics in the false negative cases. We did, however, find higher rates of false positives in Hispanics than in non-Hispanics, potentially driven by lower performance on cognitive assessments due to language and cultural barriers,^22^ which is a plausible hypothesis given that the HRS algorithms used in this study perform similarly across race/ethnicity groups. However, due to the limited number of Hispanic participants in the ADAMS study – against which the HRS algorithms were trained and validated – findings related to Hispanic ethnicity should be interpreted with caution.^6^

Among the ICD-10 codes in the DDH case definition, F03 codes – for unspecified dementia – were most frequently used uniquely to indicate dementia. In contrast, the three etiology-specific dementia codes (Alzheimer’s disease, vascular dementia, and dementia in other diseases) were rarely observed without other dementia-related codes. This may reflect the challenges of and perceived limited benefits of identifying dementia etiology in standard clinical settings.^23^ More notably, the proportion of false positives was higher in individuals with one or more of the three etiology-specific codes than in individuals with unspecified dementia in claims. We hypothesize that unspecified dementia may be more frequently coded when patients exhibit observable symptoms of cognitive decline, but lack specific indicators associated with a specific etiology. In contrast, etiology-specific dementia codes may be used more commonly in cases where other clinical evidence suggests dementia in the absence of obvious cognitive symptoms that drive dementia identification by the HRS algorithms, or when clinicians are in the process of “ruling out” specific alternate diagnoses for a given process. Finally, the R54 code for “age-related physical debility” had a higher true positive rate of 31% than all other codes in the possible category (including mild cognitive impairment, G31.84, with a true positive rate of 25%), supporting its continued use in identifying dementia in Medicare for endeavors that prioritize sensitivity over specificity. Moving forward, we recommend the exclusion of other possible category codes beyond R54 and G31.84, which have a true positive rate of under 20%.

Collectively, our findings indicate that using a code-based identification strategy such as the NORC’s DDH case definitions in Medicare claims or other administrative data substantially under-estimates population-level dementia prevalence – potentially by as much as 50% – with variation across geography and individual characteristics. These findings underscore the need for statistical modeling to calibrate claims-based surveillance and generate more accurate estimates of dementia burden at the national, state, and local level. Importantly, our examination of how different characteristics relate to over- and under-diagnosis contributes to the evidence base needed to inform such statistical calibration strategies.

The strength of this study is the utilization of the large, nationally representative HRS data to evaluate the performance of NORC’s new DDH code-based case definition in the Medicare system. Notably, this allowed for examination of geographic variation in performance, which is an essential consideration given that the case definition’s intended application is for national dementia surveillance.^4^ To preserve sample size, we used rigorous imputation methods to impute missing cognition data for the 11% of web respondents in the 2018 wave. Similarly, the use of linked MA encounters in addition to FFS claims was an important strength given the large and growing proportion of Medicare beneficiaries enrolled in MA plans.^24^ Additionally, the HRS algorithmic classifications have been validated to perform similarly across race/ethnicity groups, allowing for unbiased comparisons of mis-diagnosis by race and ethnicity.

Conversely, a limitation of using the HRS is its absence of formal dementia ascertainment, requiring us to determine dementia status – against which we evaluated the NORC code-based case definition – using probabilistic algorithms. However, because they predict dementia status based on risk indicated by collective cognitive, functional, and sociodemographic data, their bias is systematic rather than random. As such, they provide a consistent and transparent benchmark for evaluating code-based classifications. Validation against the ADAMS sub-study showed that the algorithms over-estimate dementia at the population level,^25^ implying an under-estimation of sensitivity and over-estimation of specificity, which is consistent with the findings of a recently published study evaluating NORC’s case definition against in-person research-based dementia ascertainment from the Chicago-based Rush Alzheimer’s Disease Center (RADC) cohorts.^26^

The HRS algorithms were developed and validated using ADAMS sub-study data, collected between 2000 and 2008.^6^ Potential temporal changes in the relationships between the algorithms’ predictors and dementia probability may limit the generalizability of their performance to 2018 data. Moreover, due to the small sample size of Hispanic participants in ADAMS, any conclusions pertaining to the Hispanic population should be interpreted with caution.^6^ Finally, though the HRS algorithms used in this study minimize differences in performance across race/ethnicity groups, prior work indicates that their performance across other participant characteristics can vary substantially,^25^ which should be taken into consideration when developing error-correction adjustments to diagnosed prevalence. An updated performance assessment using a larger and more current sample would be valuable. In absence of a current study with gold standard diagnoses, the Harmonized Cognitive Assessment Protocol Project (HCAP) could be leveraged as a reference point. Specifically, this HRS sub-study assigns dementia status through a multi-phase process, comparing factor scores derived from a comprehensive and in-depth neuropsychological battery and informant interview of cognitive and functional ability against a robust normative sample, and applying standard diagnostic criteria from the National Institute of Aging and Alzheimer’s Association workgroups.^27^

Consistent with our current understanding, the results of this study show that dementia is under-diagnosed in Medicare claims, with NORC’s new code-based case definition for likely-or-higher dementia achieving a sensitivity of 50%, and the definition for possible-or-higher dementia achieving a sensitivity of 59%. We also found substantial variation in performance by geography and by demographic, insurance, cognitive, and physical health characteristics, all of which have the potential to distort estimates of burden and resource allocation decisions. However, NORC’s *Dementia DataHub* (https://www.dementiadatahub.org/), which applies the case definition to estimate dementia burden at national and subnational levels, remains critical to ongoing surveillance efforts, and establishes the empirical basis from which bias-corrected and more refined estimates can be derived to inform policy and planning.

## Data Availability

All data produced in the present work are contained in the manuscript.

