## Supplemental materials for "Performance of ICD10 Code-Based Dementia Case Definition in the Health and Retirement Study"

**Appendix 1.** Imputation of cognitive survey items for 2018 HRS web respondents

To address missing data in our analysis, we employed the **MICE** (Multivariate Imputation by Chained Equations) package in R. The mice package uses an iterative approach to impute missing values by specifying a series of conditional models—one for each variable with missing data—based on the other variables in the dataset. This technique supports various types of variables (e.g., continuous, binary, categorical). The use of multiple imputation via mice allows us to minimize potential bias and loss of efficiency due to missing data, under the assumption that data are missing at random (MAR).

We generated multiple imputed datasets (typically m=10) to reflect the uncertainty inherent in the imputation process. The imputation process consisted of an initial burn-in followed by multiple iterations of chained equations, during which the values of missing data were iteratively updated.

For each imputed variable, the remaining variables in the dataset were considered for predictors. Models were fit for every combination and the top two predictors for each imputed variable were selected based on Akaike Information Criterion (AIC). Standard linear models were fit for continuous, discrete, and categorical dependent variables, probit regression models for ordinal dependent variables, and binomial regression models for dichotomous dependent variables. Predictors were only considered if they had greater than 100 non-missing values in common with the dependent variable.

Cronbach's alpha was used to measure internal consistency, indicating how closely related a set of items are as a group. Cronbach's alpha is a reliability coefficient, most commonly used to assess the reliability of scales and tests, especially those with multiple Likert questions. A higher Cronbach's alpha score (closer to 1) suggests greater internal consistency and reliability of the scale. To maintain alpha values for the mean measures of ‘namerecall’ and ‘daterecall’, additional procedures were implemented post-imputation.

For cases where all items used to calculate the mean measures were missing, the items were imputed by random selection from the set of data with entirely non-missing items, and with the same imputed mean value for the mean measure. For cases with at least one non-missing item, a similar process was conducted, however, sub-setting to cases that have the same non-missing item values. Mean measures were then calculated based on imputed item values. Cronbach alpha values were then compared to confirm internal consistency was maintained for these mean measures.

**Appendix 2.** Study stratification variable specifications.

| **Stratification variable** | **Specification** |
| --- | --- |
| Medicare Fee-for-service (FFS) vs. Medicare Advantage (MA) enrollment | Defined using the 2018 Medicare Master Beneficiary Summary File (MBSF).  Beneficiaries are defined as MA-enrolled if they have at least one month of enrollment in an MA plan during the year, and FFS-enrolled otherwise. |
| Rurality | Defined using HRS restricted cross-wave geographic information, *Beale Rural-Urban Continuum codes for 2023* variable  Urban is defined as:   1. Metro – Counties in metro areas of 1 million population and more 2. Metro - Counties in metro areas of 250,000 to 1 million population   Rural is defined as:   3. Metro - Counties in metro areas of fewer than 250,000 population 4. Nonmetro - Urban population of 20,000 or more, adjacent to a metro area 5. Nonmetro - Urban population of 20,000 or more, not adjacent to a metro area 6. Nonmetro - Urban population of 5,000 to 20,000, adjacent to a metro area 7. Nonmetro - Urban population of 5,000 to 20,000, not adjacent to a metro area 8. Nonmetro - Urban population of fewer than 5,000, adjacent to a metro area 9. Nonmetro - Urban population of fewer than 5,000, not adjacent to a metro area |
| Census division | Defined using HRS Public Use File 2018 wave, *Census Division* variable ^a^     1. New England (Connecticut; Maine; Massachusetts; New Hampshire; Rhode Island; Vermont) 2. Mid Atlantic (New Jersey; New York; Pennsylvania) 3. EN Central (Illinois; Indiana; Michigan; Ohio; Wisconsin) 4. WN Central (Iowa; Kansas; Minnesota; Missouri; Nebraska; South Dakota; South Dakota) 5. S Atlantic (Delaware; District of Columbia; Floria; Georgia; Maryland; North Carolina; South Carolina; Virginia; West Virginia) 6. ES Central (Alabama; Kentucky; Mississippi; Tennessee) 7. ES Central (Arkansas; Louisiana; Oklahoma; Texas) 8. Mountain (Arizona; Colorado; Idaho; Montana; Nevada; New Mexico; Utah; Wyoming) 9. Pacific (Alaska; California; Hawaii; Oregon; Washington) |

^a^ Census division to state mapping drawn from Census.gov (<https://www2.census.gov/geo/pdfs/maps-data/maps/reference/us_regdiv.pdf> )

**Appendix 3.** Sample Descriptive Statistics

| **Measure** | **Overall Sample, mean/% (95CI)** |
| --- | --- |
| Total N | 45,903,141 |
| *Age, %* |  |
| 65 - 69 | 28.7 (26.9 - 30.4) |
| 70 - 74 | 25.9 (24.7 - 27.1) |
| 75 - 79 | 18.7 (17.6 - 19.7) |
| 80-84 | 13.3 (12.3 - 14.3) |
| 85+ | 13.5 (12.5 - 14.4) |
| *Sex, %* |  |
| Male | 44.5 (43.6 - 45.4) |
| Female | 55.5 (54.6 - 56.4) |
| *Race/ethnicity, %* | |
| Hispanic | 8 (5.8 - 10.2) |
| Non-Hispanic Black | 8.8 (7.8 - 9.9) |
| Non-Hispanic White | 80.5 (77.9 - 83.2) |
| Other | 2.6 (2 - 3.3) |
| Greater than high school education, % | 35.1 (32.8 - 37.4) |
| Lives with partner, % | 58.5 (56.8 - 60.2) |
| *Insurance, %* |  |
| Dual-eligible | 12.5 (10.9 - 14) |
| Medicare Advantage | 41 (38.5 - 43.5) |
| *Survey response mode, %* | |
| Self respondent | 95.5 (95 - 96.1) |
| Web respondent | 11.2 (10 - 12.5) |
| *Physical health* | |
| Self-reported health, poor or fair | 27.3 (26 - 28.7) |
| 1+ ADL limitation | 19.9 (18.7 - 21.1) |
| 1+ IADL limitation | 17.8 (16.8 - 18.8) |
| High blood pressure diagnosis | 66.3 (64.7 - 67.8) |
| Diabetes diagnosis | 27.7 (26.4 - 29) |
| Died during 2018 | 1.7 (1.4 - 2.1) |
| *Cognitive health* | |
| *Self response (N=43,856,771)* | |
| Immediate word recall, mean | 5.4 (5.4 - 5.5) |
| Delayed word recall, mean | 4.5 (4.4 - 4.5) |
| Serial 7s, mean | 3.7 (3.6 - 3.7) |
| Backwards count, correct 1st attempt, % | 93.4 (92.7 - 94.1) |
| Correct date recall (4 of 4), % ^a^ | |
| Excluding web respondents | 74 (72.5 - 75.5) |
| Including web respondents | 74.4 (72.7 - 76.1) |
| Correct name recall (4 of 4), % ^a^ | |
| Excluding web respondents | 58.8 (56.8 - 60.8) |
| Including web respondents | 59.5 (52.8 - 66.2) |
| *Proxy response (N=43,856,771)* | |
| IQCODE, mean | 3.8 (3.7 - 3.9) |
| Proxy memory score, mean | 4 (3.9 - 4.1) |
| Jorm symptom count, mean | 1.5 (1.4 - 1.7) |
| *Setting, %* |  |
| Rural | 27.6 (24.3 - 31) |
| Urban | 72.4 (69 - 75.7) |
| *Census division, %* | |
| New England | 4.5 (1.3 - 7.6) |
| Middle Atlantic | 11.2 (7.8 - 14.6) |
| East North Central | 16 (13.5 - 18.5) |
| West North Central | 8.5 (6 - 11.1) |
| South Atlantic | 23.4 (20.6 - 26.2) |
| East South Central | 6.4 (4.3 - 8.6) |
| West South Central | 9.8 (7.7 - 11.8) |
| Mountain | 7 (3.8 - 10.3) |
| Pacific | 13.1 (10.4 - 15.9) |
| *Dementia status, %* | |
| Likely-or-higher (DDH) | 8.6 (7.9 - 9.4) |
| Possible-or-higher (DDH) | 13.5 (12.4 - 14.5) |
| Expert Model (HRS) | 12 (11 - 12.9) |
| LASSO Model (HRS) | 12 (11.1 - 13) |
| Modified Hurd Model (HRS) | 11.6 (10.7 - 12.5) |

Abbreviations: ADL = activities of daily living; CI = confidence interval; DDH = Dementia Datahub; GED = general education development; HRS = Health and Retirement Study; IADL = instrumental activities of daily living; IQCODE = Informant questionnaire on cognitive decline in the elderly.

^a^ These data were missing for wave 2018 web-respondents (11.2% of the sample); we used multiple imputation chained equations (MICE) to generate 10 sets of imputations for these missing data. For estimates including web respondents, point estimates were averaged across the ten datasets, and confidence intervals incorporate both within- and between-imputation variability.

Notes:

All analyses use complex variance estimation to account for the HRS survey design.

**Appendix 4.** Performance of DDH likely-or-higher dementia against HRS Expert Model dementia by census division and rural/urban setting

| **Division** | **Case Definition Performance (95% CI) ^a^** | | |
| --- | --- | --- | --- |
|  | **Sensitivity** | **Specificity** | **Accuracy** |
| *Overall* | | | |
| New England | 43 (25 - 60) | 97 (96 - 99) | 90 (86 - 94) |
| Middle Atlantic | 56 (49 - 64) | 97 (96 - 98) | 93 (91 - 94) |
| East North Central | 52 (43 - 61) | 98 (97 - 99) | 92 (90 - 93) |
| West North Central | 43 (29 - 56) | 98 (97 - 99) | 91 (88 - 94) |
| South Atlantic | 54 (47 - 60) | 96 (95 - 97) | 92 (90 - 93) |
| East South Central | 60 (43 - 78) | 98 (97 - 99) | 94 (92 - 95) |
| West South Central | 54 (43 - 64) | 94 (92 - 96) | 88 (85 - 91) |
| Mountain | 43 (32 - 55) | 97 (96 - 98) | 91 (88 - 95) |
| Pacific | 36 (26 - 45) | 97 (96 - 98) | 91 (89 - 93) |
| *p-value ^b^* | 0.06 (0.03 - 0.1) | 0.01 (0.01 - 0.01) | 0.14 (0.1 - 0.17) |
| *Rural* | | | |
| New England | 34 (0 - 71) | 96 (90 - 100) | 88 (77 - 98) |
| Middle Atlantic | 61 (49 - 73) | 100 (100 - 100) | 94 (92 - 97) |
| East North Central | 47 (35 - 60) | 99 (98 - 100) | 92 (88 - 96) |
| West North Central | 41 (32 - 50) | 98 (96 - 100) | 90 (85 - 96) |
| South Atlantic | 54 (46 - 62) | 98 (96 - 99) | 91 (88 - 93) |
| East South Central | 58 (40 - 76) | 99 (98 - 100) | 94 (91 - 97) |
| West South Central | 40 (27 - 52) | 95 (92 - 97) | 83 (77 - 89) |
| Mountain | 42 (21 - 63) | 97 (94 - 100) | 92 (86 - 98) |
| Pacific | 47 (15 - 79) | 97 (93 - 101) | 93 (86 - 99) |
| *p-value ^b^* | 0.19 (0.17 - 0.24) | 0 (0 - 0) | 0.01 (0 - 0.01) |
| *Urban* | | | |
| New England | 44 (28 - 60) | 98 (95 - 100) | 90 (86 - 95) |
| Middle Atlantic | 55 (48 - 62) | 97 (95 - 98) | 93 (91 - 94) |
| East North Central | 55 (45 - 66) | 97 (96 - 99) | 92 (91 - 93) |
| West North Central | 44 (18 - 71) | 98 (96 - 100) | 92 (90 - 95) |
| South Atlantic | 53 (44 - 63) | 96 (95 - 97) | 92 (90 - 94) |
| East South Central | 62 (38 - 86) | 98 (96 - 100) | 93 (91 - 96) |
| West South Central | 65 (54 - 77) | 94 (92 - 96) | 90 (88 - 93) |
| Mountain | 44 (32 - 56) | 97 (96 - 98) | 91 (88 - 95) |
| Pacific | 35 (27 - 44) | 97 (96 - 98) | 91 (89 - 92) |
| *p-value ^b^* | 0.01 (0 - 0.02) | 0.13 (0.07 - 0.13) | 0.71 (0.64 - 0.84) |

Abbreviations: DDH = Dementia Datahub; HRS = Health and Retirement Study.

^a^ Values in paratheses represent 95% CI except for p-values, for which values in parentheses represent minimum and maximum of estimates obtained across the 10 imputed datasets.

^b^ Reported p-values represent the median (minimum - maximum) of estimates obtained across the 10 imputed datasets. P-values indicate significance in differences between census divisions.

Notes: A subset of self-report cognitive data were missing for wave 2018 web-respondents (11.2% of the sample); we used multiple imputation chained equations (MICE) to generate 10 sets of imputations for these missing data. Point estimates were averaged across the ten datasets, and confidence intervals incorporate both within- and between-imputation variability.

All analyses use complex variance estimation to account for the HRS survey design.

**Appendix 5.** Probability of dementia estimated using the HRS Expert Model for participants classified as highly likely, likely-or-higher, and possible-or-higher using DDH case definitions.

| **Classification** | **N (weighted)** | **Predicted Dementia Probability using Expert Model ^a^** | | | | | |
| --- | --- | --- | --- | --- | --- | --- | --- |
|  |  | **Mean (95% CI)** | **Minimum** | **25th percentile** | **Median** | **75th percentile** | **Maximum** |
| *DDH highly likely dementia* | | | | | | | |
| True Positive | 2,391,163 | 0.817 (0.797 - 0.837) | 0.271 | 0.697 | 0.920 | 0.984 | 1.000 |
| False Negative | 3,095,570 | 0.619 (0.598 - 0.641) | 0.270 | 0.421 | 0.610 | 0.830 | 0.998 |
| True Negative | 9,663,108 | 0.024 (0.022 - 0.025) | 0.000 | 0.000 | 0.000 | 0.018 | 0.452 |
| False Positive | 753,300 | 0.082 (0.066 - 0.098) | 0.000 | 0.007 | 0.040 | 0.145 | 0.423 |
| *DDH likely-or-higher dementia* | | | | | | | |
| True Positive | 2,732,451 | 0.802 (0.782 - 0.822) | 0.271 | 0.673 | 0.890 | 0.977 | 1.000 |
| False Negative | 2,754,282 | 0.610 (0.588 - 0.631) | 0.270 | 0.414 | 0.590 | 0.808 | 0.998 |
| True Negative | 39,181,666 | 0.023 (0.022 - 0.025) | 0.000 | 0.000 | 0.000 | 0.017 | 0.452 |
| False Positive | 1,234,742 | 0.074 (0.061 - 0.087) | 0.000 | 0.005 | 0.030 | 0.122 | 0.423 |
| *DDH possible-or-higher dementia* | | | | | | | |
| True Positive | 3,208,002 | 0.779 (0.761 - 0.798) | 0.271 | 0.614 | 0.860 | 0.972 | 1.000 |
| False Negative | 2,278,731 | 0.601 (0.577 - 0.625) | 0.270 | 0.410 | 0.580 | 0.790 | 0.996 |
| True Negative | 37,443,877 | 0.022 (0.021 - 0.024) | 0.000 | 0.000 | 0.000 | 0.015 | 0.452 |
| False Positive | 2,972,531 | 0.059 (0.051 - 0.066) | 0.000 | 0.004 | 0.020 | 0.089 | 0.423 |

Abbreviations: DDH = Dementia Datahub; HRS = Health and Retirement Study.

^a^ Cells that show 0.000 are near zeroes rather than true zeroes.
Notes: A subset of self-report cognitive data were missing for wave 2018 web-respondents (11.2% of the sample); we used multiple imputation chained equations (MICE) to generate 10 sets of imputations for these missing data. Point estimates were averaged across the ten datasets, and confidence intervals incorporate both within- and between-imputation variability.

All analyses use complex variance estimation to account for the HRS survey design.

**Appendix 6.** Performance of DDH possible-or-higher dementia against HRS Expert Model dementia by census division and rural/urban Setting

| **Division** | **Case Definition Performance (95% CI) ^a^** | | |
| --- | --- | --- | --- |
|  | **Sensitivity** | **Specificity** | **Accuracy** |
| *Overall* | | | |
| New England | 48 (30 - 66) | 95 (92 - 98) | 89 (84 - 93) |
| Middle Atlantic | 65 (57 - 73) | 93 (91 - 95) | 90 (88 - 92) |
| East North Central | 62 (55 - 69) | 93 (91 - 96) | 89 (87 - 91) |
| West North Central | 52 (42 - 63) | 92 (90 - 94) | 87 (84 - 91) |
| South Atlantic | 60 (54 - 65) | 92 (91 - 94) | 89 (87 - 91) |
| East South Central | 72 (58 - 86) | 94 (92 - 96) | 91 (89 - 94) |
| West South Central | 63 (45 - 80) | 90 (88 - 93) | 86 (82 - 90) |
| Mountain | 56 (45 - 66) | 94 (92 - 97) | 90 (86 - 94) |
| Pacific | 45 (35 - 55) | 92 (89 - 95) | 87 (84 - 90) |
| *p-value ^b^* | 0.09 (0.07 - 0.11) | 0.37 (0.32 - 0.39) | 0.47 (0.46 - 0.51) |
| *Rural* | | | |
| New England | 34 (0 - 71) | 96 (90 - 100) | 88 (77 - 98) |
| Middle Atlantic | 70 (56 - 84) | 93 (89 - 97) | 90 (86 - 94) |
| East North Central | 57 (46 - 69) | 95 (92 - 98) | 90 (87 - 92) |
| West North Central | 48 (38 - 58) | 92 (88 - 96) | 86 (80 - 93) |
| South Atlantic | 57 (49 - 64) | 92 (88 - 95) | 86 (82 - 90) |
| East South Central | 62 (46 - 77) | 95 (91 - 98) | 91 (86 - 95) |
| West South Central | 52 (21 - 83) | 91 (86 - 96) | 83 (74 - 91) |
| Mountain | 55 (39 - 70) | 94 (91 - 97) | 90 (86 - 95) |
| Pacific | 63 (14 - 111) | 97 (93 - 101) | 94 (88 - 99) |
| *p-value ^b^* | 0.36 (0.23 - 0.43) | 0.52 (0.51 - 0.53) | 0.1 (0.09 - 0.13) |
| *Urban* | | | |
| New England | 50 (34 - 66) | 95 (91 - 99) | 89 (84 - 93) |
| Middle Atlantic | 64 (57 - 71) | 93 (91 - 96) | 90 (88 - 93) |
| East North Central | 65 (57 - 74) | 92 (89 - 95) | 89 (87 - 91) |
| West North Central | 57 (41 - 73) | 92 (90 - 94) | 88 (86 - 90) |
| South Atlantic | 62 (53 - 70) | 93 (91 - 94) | 90 (88 - 92) |
| East South Central | 77 (60 - 94) | 94 (90 - 97) | 91 (88 - 95) |
| West South Central | 71 (59 - 84) | 90 (87 - 93) | 88 (84 - 91) |
| Mountain | 56 (43 - 70) | 94 (91 - 97) | 90 (85 - 94) |
| Pacific | 44 (36 - 51) | 91 (88 - 94) | 86 (83 - 89) |
| *p-value ^b^* | 0 (0 - 0.01) | 0.75 (0.69 - 0.76) | 0.56 (0.48 - 0.61) |

Abbreviations: DDH = Dementia Datahub; HRS = Health and Retirement Study.

^a^ Values in paratheses represent 95% CI except for p-values, for which values in parentheses represent minimum and maximum of estimates obtained across the 10 imputed datasets.

^b^ Reported p-values represent the median (minimum - maximum) of estimates obtained across the 10 imputed datasets. P-values indicate significance in differences between census divisions.

Notes: A subset of self-report cognitive data were missing for wave 2018 web-respondents (11.2% of the sample); we used multiple imputation chained equations (MICE) to generate 10 sets of imputations for these missing data. Point estimates were averaged across the ten datasets, and confidence intervals incorporate both within- and between-imputation variability.

All analyses use complex variance estimation to account for the HRS survey design.

**Appendix 7.** Performance of DDH likely-or-higher dementia against HRS dementia, sensitivity analyses

| **Sample** | **Performance Metrics (95% CI) ^a^** | | | **Classification Distribution (95% CI) ^a^** | | | |
| --- | --- | --- | --- | --- | --- | --- | --- |
|  | **Sensitivity** | **Specificity** | **Accuracy** | **True positive** | **False negative** | **True negative** | **False positive** |
| *LASSO Model* | | | | | | | |
| **Overall** | 47 (44 - 51) | 97 (97 - 98) | 91 (90 - 92) | 5.7 (5.1 - 6.4) | 6.3 (5.7 - 6.9) | 85.5 (84.4 - 86.5) | 2.5 (2.1 - 2.9) |
| **Insurance type** | | | | | | | |
| FFS | 50 (46 - 54) | 97 (97 - 98) | 92 (91 - 93) | 5.8 (5 - 6.7) | 5.8 (5.1 - 6.6) | 85.8 (84.4 - 87.2) | 2.6 (2.1 - 3.1) |
| MA | 44 (39 - 48) | 97 (96 - 98) | 91 (89 - 92) | 5.5 (4.8 - 6.2) | 7 (6 - 8.1) | 85 (83.5 - 86.6) | 2.4 (1.8 - 3.1) |
| p-value ^b^ | 0.02 (0.02 - 0.04) | 0.78 (0.69 - 0.92) | 0.22 (0.17 - 0.27) | 0.13 (0.11 - 0.2) | | | |
| **Setting** | | | | | | | |
| Rural | 45 (40 - 50) | 98 (97 - 98) | 90 (89 - 92) | 6.4 (5.1 - 7.6) | 7.7 (6.1 - 9.3) | 83.9 (81.5 - 86.3) | 2 (1.4 - 2.6) |
| Urban | 49 (44 - 53) | 97 (96 - 97) | 91 (91 - 92) | 5.5 (4.7 - 6.2) | 5.8 (5.1 - 6.5) | 86 (84.8 - 87.2) | 2.7 (2.2 - 3.2) |
| p-value ^b^ | 0.3 (0.24 - 0.4) | 0.11 (0.1 - 0.14) | 0.2 (0.17 - 0.28) | 0.11 (0.08 - 0.14) | | | |
| *Modified Hurd Model* | | | | | | | |
| **Overall** | 51 (48 - 54) | 97 (96 - 97) | 92 (91 - 92) | 5.9 (5.3 - 6.5) | 5.7 (5.2 - 6.2) | 85.7 (84.7 - 86.7) | 2.7 (2.3 - 3.1) |
| **Insurance type** | | | | | | | |
| FFS | 53 (49 - 58) | 97 (96 - 98) | 92 (91 - 93) | 6.1 (5.2 - 6.9) | 5.3 (4.7 - 5.9) | 85.9 (84.5 - 87.3) | 2.7 (2.2 - 3.2) |
| MA | 48 (43 - 52) | 97 (96 - 98) | 91 (90 - 92) | 5.7 (4.9 - 6.4) | 6.3 (5.3 - 7.2) | 85.4 (83.8 - 86.9) | 2.7 (2 - 3.4) |
| p-value ^b^ | 0.04 (0.03 - 0.05) | 0.97 (0.87 - 1) | 0.23 (0.2 - 0.26) | 0.22 (0.19 - 0.27) | | | |
| **Setting** | | | | | | | |
| Rural | 50 (46 - 53) | 97 (97 - 98) | 91 (90 - 92) | 6.5 (5.4 - 7.7) | 6.7 (5.7 - 7.6) | 84.6 (82.8 - 86.4) | 2.3 (1.6 - 2.9) |
| Urban | 52 (47 - 56) | 97 (96 - 97) | 92 (91 - 93) | 5.7 (4.9 - 6.4) | 5.3 (4.7 - 5.9) | 86.1 (84.9 - 87.3) | 2.9 (2.4 - 3.4) |
| p-value ^b^ | 0.41 (0.36 - 0.52) | 0.17 (0.16 - 0.2) | 0.29 (0.23 - 0.36) | 0.09 (0.06 - 0.15) | | | |
| *Expert Model, excluding web respondents* | | | | | | | |
| **Overall** | 50 (47 - 54) | 97 (96 - 97) | 91 (90 - 91) | 6.7 (5.9 - 7.4) | 6.6 (5.9 - 7.2) | 83.9 (82.8 - 85) | 2.8 (2.4 - 3.3) |
| **Insurance type** | | | | | | | |
| FFS | 53 (49 - 58) | 97 (96 - 98) | 91 (90 - 92) | 7 (6 - 8) | 6.1 (5.3 - 6.9) | 84.1 (82.4 - 85.8) | 2.8 (2.2 - 3.4) |
| MA | 46 (42 - 51) | 97 (96 - 98) | 90 (89 - 91) | 6.2 (5.4 - 7.1) | 7.2 (6.2 - 8.2) | 83.6 (82.1 - 85.1) | 2.9 (2.1 - 3.7) |
| p-value | 0.01 | 0.74 | 0.14 | 0.05 | | | |
| **Setting** | | | | | | | |
| Rural | 48 (43 - 53) | 98 (97 - 98) | 90 (88 - 92) | 7.5 (6.2 - 8.9) | 8.1 (6.2 - 9.9) | 82.4 (79.7 - 85.1) | 2 (1.3 - 2.7) |
| Urban | 51 (47 - 56) | 96 (96 - 97) | 91 (90 - 92) | 6.3 (5.4 - 7.2) | 6 (5.4 - 6.6) | 84.5 (83.4 - 85.7) | 3.2 (2.6 - 3.8) |
| p-value | 0.30 | 0.03 | 0.34 | 0.04 | | | |

Abbreviations: CI = confidence interval; DDH = Dementia Datahub; FFS = fee-for-service Medicare; HRS = Health and Retirement Study; MA = Medicare Advantage.

^a^ Values in paratheses represent 95% CI except for p-values, for which values in parentheses represent minimum and maximum of estimates obtained across the 10 imputed datasets.

^b^ Reported p-values represent the median (minimum - maximum) of estimates obtained across the 10 imputed datasets. . P-values indicate significance in differences between subgroups (FFS vs. MA; Rural vs. Urban).

Notes: A subset of self-report cognitive data were missing for wave 2018 web-respondents (11.2% of the sample); we used multiple imputation chained equations (MICE) to generate 10 sets of imputations for these missing data. Point estimates were averaged across the ten datasets, and confidence intervals incorporate both within- and between-imputation variability. PPV/NPV are omitted because they vary with subgroup prevalence and are not comparable across subgroup populations.

All analyses use complex variance estimation to account for the HRS survey design.

**Appendix 8.** Performance of DDH likely-or-higher dementia against HRS LASSO Model dementia by census division and rural/urban Setting

| **Division** | **Case Definition Performance (95% CI) ^a^** | | |
| --- | --- | --- | --- |
|  | **Sensitivity** | **Specificity** | **Accuracy** |
| *Overall* | | | |
| New England | 46 (29 - 63) | 98 (97 - 100) | 92 (90 - 94) |
| Middle Atlantic | 50 (43 - 58) | 97 (96 - 98) | 92 (90 - 94) |
| East North Central | 49 (40 - 57) | 98 (97 - 99) | 91 (89 - 93) |
| West North Central | 39 (25 - 52) | 98 (96 - 99) | 91 (89 - 93) |
| South Atlantic | 51 (44 - 58) | 96 (95 - 97) | 91 (89 - 93) |
| East South Central | 56 (38 - 73) | 98 (97 - 100) | 93 (91 - 95) |
| West South Central | 53 (43 - 63) | 95 (94 - 97) | 89 (86 - 92) |
| Mountain | 42 (32 - 52) | 97 (96 - 99) | 92 (89 - 96) |
| Pacific | 35 (25 - 46) | 97 (96 - 98) | 91 (88 - 93) |
| *p-value ^b^* | 0.3 (0.08 - 0.37) | 0.06 (0.03 - 0.06) | 0.61 (0.58 - 0.68) |
| *Rural* | | | |
| New England | 65 (34 - 96) | 98 (95 - 101) | 95 (90 - 100) |
| Middle Atlantic | 46 (43 - 50) | 100 (100 - 100) | 92 (86 - 97) |
| East North Central | 44 (34 - 54) | 99 (98 - 99) | 91 (87 - 94) |
| West North Central | 36 (23 - 49) | 97 (96 - 98) | 90 (87 - 92) |
| South Atlantic | 53 (44 - 62) | 97 (96 - 99) | 90 (87 - 94) |
| East South Central | 59 (47 - 72) | 99 (98 - 100) | 94 (91 - 97) |
| West South Central | 37 (26 - 48) | 95 (92 - 98) | 84 (77 - 90) |
| Mountain | 36 (10 - 63) | 97 (94 - 100) | 91 (84 - 98) |
| Pacific | 38 (0 - 77) | 97 (93 - 101) | 92 (86 - 98) |
| *p-value ^b^* | 0.24 (0.23 - 0.27) | 0 (0 - 0) | 0.04 (0.02 - 0.06) |
| *Urban* | | | |
| New England | 44 (23 - 64) | 99 (97 - 100) | 92 (89 - 94) |
| Middle Atlantic | 51 (42 - 60) | 97 (96 - 98) | 92 (89 - 95) |
| East North Central | 52 (39 - 65) | 97 (96 - 99) | 91 (88 - 94) |
| West North Central | 41 (20 - 63) | 98 (96 - 100) | 92 (90 - 94) |
| South Atlantic | 50 (41 - 59) | 96 (95 - 97) | 92 (90 - 93) |
| East South Central | 54 (29 - 79) | 98 (96 - 100) | 92 (89 - 95) |
| West South Central | 65 (54 - 76) | 95 (94 - 97) | 91 (89 - 94) |
| Mountain | 45 (36 - 54) | 97 (96 - 99) | 92 (89 - 96) |
| Pacific | 35 (25 - 45) | 97 (96 - 99) | 90 (88 - 93) |
| *p-value ^b^* | 0.04 (0.01 - 0.14) | 0.31 (0.15 - 0.31) | 0.99 (0.97 - 1) |

Abbreviations: DDH = Dementia Datahub; HRS = Health and Retirement Study.

^a^ Values in paratheses represent 95% CI except for p-values, for which values in parentheses represent minimum and maximum of estimates obtained across the 10 imputed datasets.

^b^ Reported p-values represent the median (minimum - maximum) of estimates obtained across the 10 imputed datasets. P-values indicate significance in differences between census divisions.

Notes: A subset of self-report cognitive data were missing for wave 2018 web-respondents (11.2% of the sample); we used multiple imputation chained equations (MICE) to generate 10 sets of imputations for these missing data. Point estimates were averaged across the ten datasets, and confidence intervals incorporate both within- and between-imputation variability.

All analyses use complex variance estimation to account for the HRS survey design.

**Appendix 9.** Performance of DDH likely-or-higher dementia against HRS Modified-Hurd Model dementia by census division and rural/urban setting

| **Division** | **Case Definition Performance (95% CI) ^a^** | | |
| --- | --- | --- | --- |
|  | **Sensitivity** | **Specificity** | **Accuracy** |
| *Overall* | | | |
| New England | 49 (28 - 71) | 97 (96 - 98) | 92 (88 - 95) |
| Middle Atlantic | 53 (49 - 58) | 97 (96 - 98) | 92 (91 - 94) |
| East North Central | 54 (47 - 62) | 98 (97 - 99) | 92 (91 - 94) |
| West North Central | 43 (29 - 57) | 97 (96 - 99) | 92 (90 - 93) |
| South Atlantic | 54 (47 - 62) | 96 (95 - 97) | 92 (90 - 94) |
| East South Central | 60 (46 - 74) | 98 (96 - 100) | 93 (91 - 96) |
| West South Central | 51 (44 - 59) | 94 (92 - 96) | 88 (85 - 90) |
| Mountain | 53 (38 - 68) | 98 (97 - 99) | 93 (90 - 96) |
| Pacific | 39 (31 - 46) | 98 (97 - 99) | 91 (89 - 93) |
| *p-value ^b^* | 0.13 (0.06 - 0.2) | 0.02 (0.01 - 0.02) | 0.16 (0.12 - 0.17) |
| *Rural* | | | |
| New England | 35 (-7 - 78) | 94 (88 - 100) | 89 (83 - 96) |
| Middle Atlantic | 54 (50 - 59) | 100 (100 - 100) | 93 (89 - 97) |
| East North Central | 51 (44 - 59) | 98 (97 - 99) | 92 (90 - 95) |
| West North Central | 43 (32 - 55) | 97 (96 - 98) | 91 (89 - 93) |
| South Atlantic | 53 (45 - 60) | 97 (96 - 99) | 90 (87 - 93) |
| East South Central | 70 (52 - 89) | 99 (98 - 100) | 96 (93 - 99) |
| West South Central | 38 (30 - 45) | 95 (92 - 97) | 83 (79 - 88) |
| Mountain | 58 (36 - 81) | 97 (94 - 100) | 94 (89 - 99) |
| Pacific | 42 (7 - 77) | 97 (93 - 100) | 91 (83 - 99) |
| *p-value ^b^* | 0.08 (0.06 - 0.1) | 0 (0 - 0) | 0.01 (0.01 - 0.01) |
| *Urban* | | | |
| New England | 51 (30 - 72) | 98 (96 - 99) | 92 (88 - 96) |
| Middle Atlantic | 53 (48 - 58) | 96 (95 - 97) | 92 (91 - 94) |
| East North Central | 56 (46 - 67) | 98 (96 - 99) | 92 (90 - 94) |
| West North Central | 42 (19 - 66) | 98 (96 - 100) | 92 (90 - 95) |
| South Atlantic | 55 (45 - 66) | 96 (95 - 97) | 92 (90 - 94) |
| East South Central | 55 (37 - 74) | 97 (95 - 100) | 92 (88 - 95) |
| West South Central | 61 (50 - 72) | 94 (91 - 96) | 89 (86 - 93) |
| Mountain | 51 (32 - 70) | 98 (97 - 99) | 93 (90 - 96) |
| Pacific | 39 (32 - 45) | 98 (97 - 99) | 91 (89 - 93) |
| *p-value ^b^* | 0.08 (0.03 - 0.13) | 0.06 (0.05 - 0.06) | 0.89 (0.84 - 0.91) |

Abbreviations: DDH = Dementia Datahub; HRS = Health and Retirement Study.

^a^ Values in paratheses represent 95% CI except for p-values, for which values in parentheses represent minimum and maximum of estimates obtained across the 10 imputed datasets.

^b^ Reported p-values represent the median (minimum - maximum) of estimates obtained across the 10 imputed datasets. P-values indicate significance in differences between census divisions.

Notes: A subset of self-report cognitive data were missing for wave 2018 web-respondents (11.2% of the sample); we used multiple imputation chained equations (MICE) to generate 10 sets of imputations for these missing data. Point estimates were averaged across the ten datasets, and confidence intervals incorporate both within- and between-imputation variability.

All analyses use complex variance estimation to account for the HRS survey design.

**Appendix 10.** Performance of DDH likely-or-higher dementia against HRS Expert Model dementia by census division and rural/urban setting, excluding web respondents

| **Division** | **Case Definition Performance (95% CI)** | | |
| --- | --- | --- | --- |
|  | **Sensitivity** | **Specificity** | **Accuracy** |
| *Overall* | | | |
| New England | 44 (25 - 63) | 97 (95 - 99) | 89 (85 - 93) |
| Middle Atlantic | 56 (49 - 64) | 97 (95 - 98) | 92 (91 - 94) |
| East North Central | 52 (43 - 62) | 98 (96 - 99) | 91 (89 - 93) |
| West North Central | 44 (30 - 58) | 98 (96 - 99) | 91 (88 - 94) |
| South Atlantic | 54 (48 - 60) | 96 (95 - 97) | 91 (89 - 92) |
| East South Central | 60 (42 - 78) | 98 (97 - 100) | 93 (91 - 95) |
| West South Central | 54 (43 - 65) | 94 (92 - 96) | 88 (85 - 91) |
| Mountain | 45 (33 - 58) | 97 (96 - 99) | 91 (88 - 95) |
| Pacific | 36 (27 - 46) | 97 (96 - 98) | 90 (88 - 92) |
| *p-value ^a^* | 0.08 | 0.04 | 0.19 |
| *Rural* | | | |
| New England | 37 (0 - 78) | 95 (90 - 100) | 88 (78 - 98) |
| Middle Atlantic | 61 (49 - 73) | 100 (100 - 100) | 94 (91 - 96) |
| East North Central | 48 (34 - 61) | 98 (97 - 100) | 91 (86 - 95) |
| West North Central | 43 (35 - 52) | 98 (96 - 100) | 90 (85 - 95) |
| South Atlantic | 54 (46 - 62) | 97 (96 - 99) | 90 (86 - 93) |
| East South Central | 58 (40 - 76) | 99 (98 - 100) | 94 (91 - 97) |
| West South Central | 40 (27 - 52) | 95 (92 - 98) | 83 (75 - 90) |
| Mountain | 42 (20 - 64) | 97 (94 - 100) | 91 (85 - 98) |
| Pacific | 50 (21 - 79) | 97 (92 - 100) | 92 (84 - 100) |
| *p-value ^a^* | 0.23 | 0.00 | 0.03 |
| *Urban* | | | |
| New England | 45 (28 - 63) | 97 (94 - 100) | 89 (84 - 94) |
| Middle Atlantic | 55 (48 - 63) | 96 (95 - 98) | 92 (90 - 94) |
| East North Central | 56 (45 - 67) | 97 (95 - 99) | 91 (90 - 92) |
| West North Central | 45 (18 - 71) | 98 (95 - 100) | 91 (88 - 94) |
| South Atlantic | 54 (44 - 63) | 95 (94 - 97) | 91 (89 - 93) |
| East South Central | 62 (38 - 86) | 98 (96 - 100) | 92 (90 - 95) |
| West South Central | 66 (54 - 78) | 94 (91 - 96) | 90 (87 - 93) |
| Mountain | 47 (34 - 59) | 97 (96 - 99) | 91 (88 - 94) |
| Pacific | 35 (27 - 44) | 97 (96 - 98) | 90 (87 - 92) |
| *p-value ^a^* | 0.01 | 0.17 | 0.78 |

Abbreviations: DDH = Dementia Datahub; HRS = Health and Retirement Study.

^a^ P-values indicate significance in differences between census divisions.

Notes: A subset of self-report cognitive data were missing for wave 2018 web-respondents (11.2% of the sample); they are excluded from this analysis.

All analyses use complex variance estimation to account for the HRS survey design.

**Appendix 11.** Probability of dementia estimated using the HRS LASSO Model for participants classified as highly likely, likely-or-higher, and possible-or-higher using DDH case definitions.

| **Classification** | **N** | **Predicted Dementia Probability ^a^** | | | | | |
| --- | --- | --- | --- | --- | --- | --- | --- |
|  |  | **Mean (95% CI)** | **Minimum** | **25th percentile** | **Median** | **75th percentile** | **Maximum** |
| *DDH highly likely dementia* | | | | | | | |
| True Positive | 2,498,339 | 0.807 (0.786 - 0.828) | 0.200 | 0.690 | 0.920 | 0.988 | 1.000 |
| False Negative | 2,771,150 | 0.534 (0.514 - 0.554) | 0.191 | 0.334 | 0.490 | 0.710 | 0.998 |
| True Negative | 37,388,465 | 0.033 (0.031 - 0.035) | 0.000 | 0.004 | 0.010 | 0.039 | 0.332 |
| False Positive | 1,096,389 | 0.084 (0.07 - 0.099) | 0.004 | 0.025 | 0.050 | 0.140 | 0.320 |
| *DDH likely-or-higher dementia* | | | | | | | |
| True Positive | 2,961,323 | 0.79 (0.769 - 0.812) | 0.200 | 0.642 | 0.890 | 0.983 | 1.000 |
| False Negative | 2,308,165 | 0.519 (0.499 - 0.54) | 0.191 | 0.323 | 0.460 | 0.692 | 0.998 |
| True Negative | 35,776,997 | 0.033 (0.031 - 0.034) | 0.000 | 0.004 | 0.010 | 0.038 | 0.332 |
| False Positive | 2,707,857 | 0.082 (0.07 - 0.094) | 0.001 | 0.021 | 0.050 | 0.136 | 0.320 |
| *DDH possible-or-higher dementia* | | | | | | | |
| True Positive | 2,197,881 | 0.754 (0.733 - 0.774) | 0.192 | 0.553 | 0.840 | 0.977 | 1.000 |
| False Negative | 3,071,608 | 0.512 (0.491 - 0.533) | 0.191 | 0.328 | 0.450 | 0.681 | 0.998 |
| True Negative | 37,834,531 | 0.031 (0.03 - 0.033) | 0.000 | 0.004 | 0.010 | 0.036 | 0.332 |
| False Positive | 650,323 | 0.069 (0.062 - 0.076) | 0.000 | 0.015 | 0.040 | 0.111 | 0.320 |

Abbreviations: DDH = Dementia Datahub; HRS = Health and Retirement Study.

^a^ Cells that show 0.000 are near zeroes rather than true zeroes.
Notes: A subset of self-report cognitive data were missing for wave 2018 web-respondents (11.2% of the sample); we used multiple imputation chained equations (MICE) to generate 10 sets of imputations for these missing data. Point estimates were averaged across the ten datasets, and confidence intervals incorporate both within- and between-imputation variability.

All analyses use complex variance estimation to account for the HRS survey design.

**Appendix 12.** Probability of dementia estimated using the HRS Modified-Hurd Model for participants classified as highly likely, likely-or-higher, and possible-or-higher using DDH case definitions.

| **Classification** | **N** | **Predicted Dementia Probability ^a^** | | | | | |
| --- | --- | --- | --- | --- | --- | --- | --- |
|  |  | **Mean (95% CI)** | **Minimum** | **25th percentile** | **Median** | **75th percentile** | **Maximum** |
| *DDH highly likely dementia* | | | | | | | |
| True Positive | 2,498,339 | 0.773 (0.749 - 0.798) | 0.191 | 0.571 | 0.910 | 1.000 | 1.000 |
| False Negative | 2,771,150 | 0.478 (0.452 - 0.503) | 0.190 | 0.274 | 0.400 | 0.653 | 1.000 |
| True Negative | 37,388,465 | 0.02 (0.019 - 0.021) | 0.000 | 0.001 | 0.000 | 0.021 | 0.267 |
| False Positive | 1,096,389 | 0.066 (0.055 - 0.078) | 0.000 | 0.010 | 0.050 | 0.109 | 0.266 |
| *DDH likely-or-higher dementia* | | | | | | | |
| True Positive | 2,961,323 | 0.747 (0.723 - 0.771) | 0.191 | 0.523 | 0.830 | 0.999 | 1.000 |
| False Negative | 2,308,165 | 0.465 (0.439 - 0.491) | 0.190 | 0.269 | 0.390 | 0.607 | 1.000 |
| True Negative | 35,776,997 | 0.02 (0.019 - 0.021) | 0.000 | 0.001 | 0.000 | 0.020 | 0.267 |
| False Positive | 2,707,857 | 0.059 (0.05 - 0.068) | 0.000 | 0.009 | 0.040 | 0.095 | 0.266 |
| *DDH possible-or-higher dementia* | | | | | | | |
| True Positive | 2,197,881 | 0.712 (0.692 - 0.732) | 0.191 | 0.441 | 0.780 | 0.997 | 1.000 |
| False Negative | 3,071,608 | 0.454 (0.426 - 0.482) | 0.190 | 0.271 | 0.380 | 0.572 | 1.000 |
| True Negative | 37,834,531 | 0.019 (0.018 - 0.02) | 0.000 | 0.000 | 0.000 | 0.019 | 0.259 |
| False Positive | 650,323 | 0.05 (0.045 - 0.056) | 0.000 | 0.004 | 0.030 | 0.081 | 0.267 |

Abbreviations: DDH = Dementia Datahub; HRS = Health and Retirement Study.

^a^ Cells that show 0.000 are near zeroes rather than true zeroes.
Notes: A subset of self-report cognitive data were missing for wave 2018 web-respondents (11.2% of the sample); we used multiple imputation chained equations (MICE) to generate 10 sets of imputations for these missing data. Point estimates were averaged across the ten datasets, and confidence intervals incorporate both within- and between-imputation variability.

All analyses use complex variance estimation to account for the HRS survey design.

**Appendix 13.** Probability of dementia estimated using the HRS Expert Model for participants classified as highly likely, likely-or-higher, and possible-or-higher using DDH case definitions, excluding web respondents

| **Classification** | **N** | **Predicted Dementia Probability ^a^** | | | | | |
| --- | --- | --- | --- | --- | --- | --- | --- |
|  |  | **Mean (95% CI)** | **Minimum** | **25th percentile** | **Median** | **75th percentile** | **Maximum** |
| *DDH highly likely dementia* | | | | | | | |
| True Positive | 2,498,339 | 0.819 (0.799 - 0.839) | 0.271 | 0.699 | 0.920 | 0.984 | 1.000 |
| False Negative | 2,771,150 | 0.622 (0.6 - 0.644) | 0.270 | 0.423 | 0.610 | 0.833 | 0.998 |
| True Negative | 37,388,465 | 0.026 (0.024 - 0.027) | 0.000 | 0.001 | 0.000 | 0.021 | 0.452 |
| False Positive | 1,096,389 | 0.083 (0.066 - 0.1) | 0.000 | 0.007 | 0.040 | 0.148 | 0.423 |
| *DDH likely-or-higher dementia* | | | | | | | |
| True Positive | 2,961,323 | 0.804 (0.784 - 0.823) | 0.271 | 0.674 | 0.890 | 0.978 | 1.000 |
| False Negative | 2,308,165 | 0.613 (0.59 - 0.635) | 0.270 | 0.415 | 0.590 | 0.818 | 0.998 |
| True Negative | 35,776,997 | 0.025 (0.023 - 0.027) | 0.000 | 0.001 | 0.000 | 0.020 | 0.452 |
| False Positive | 2,707,857 | 0.075 (0.061 - 0.089) | 0.000 | 0.005 | 0.030 | 0.127 | 0.423 |
| *DDH possible-or-higher dementia* | | | | | | | |
| True Positive | 2,197,881 | 0.781 (0.763 - 0.8) | 0.271 | 0.618 | 0.860 | 0.972 | 1.000 |
| False Negative | 3,071,608 | 0.604 (0.58 - 0.629) | 0.270 | 0.412 | 0.580 | 0.795 | 0.996 |
| True Negative | 37,834,531 | 0.024 (0.022 - 0.026) | 0.000 | 0.000 | 0.000 | 0.018 | 0.452 |
| False Positive | 650,323 | 0.06 (0.052 - 0.067) | 0.000 | 0.005 | 0.020 | 0.091 | 0.423 |

Abbreviations: DDH = Dementia Datahub; HRS = Health and Retirement Study.

^a^ Cells that show 0.000 are near zeroes rather than true zeroes.
Notes: A subset of self-report cognitive data were missing for wave 2018 web-respondents (11.2% of the sample); they were excluded from this analysis.

All analyses use complex variance estimation to account for the HRS survey design.

**Appendix 14.** Sample Descriptive Statistics, overall and by HRS Expert Model and DDH possible-or-higher classifications

| **Measure** | **Overall sample, mean/% (95CI)** | **HRS vs. DDH classification, Mean/% (95% CI)** | | | | | |
| --- | --- | --- | --- | --- | --- | --- | --- |
|  |  | **True positive**  **(TP)** | **False negative (FN)** | **TP vs. FN p-value ^a^** | **True negative (TN)** | **False positive (FP)** | **TP vs. FN p-value ^a^** |
| Total N | 45,903,141 | 3,208,002 | 2,278,731 |  | 37,443,878 | 2,972,531 |  |
| *Age, %* | | | | | | | |
| 65 - 69 | 28.7 (26.9 - 30.4) | 2.5 (0.6 - 4.4) | 0.7 (0 - 1.3) | ** | 33.5 (31.7 - 35.2) | 17.9 (12.6 - 23.2) | *** |
| 70 - 74 | 25.9 (24.7 - 27.1) | 6.7 (4.2 - 9.2) | 6.9 (3.4 - 10.4) |  | 29.1 (27.7 - 30.5) | 21 (16.5 - 25.4) |  |
| 75 - 79 | 18.7 (17.6 - 19.7) | 12.4 (9.4 - 15.4) | 16.5 (13.2 - 19.9) |  | 18.9 (17.9 - 19.9) | 24 (19.2 - 28.8) |  |
| 80-84 | 13.3 (12.3 - 14.3) | 23.4 (20.1 - 26.7) | 25.7 (22.6 - 28.8) |  | 11.2 (10.2 - 12.3) | 19 (14.9 - 23) |  |
| 85+ | 13.5 (12.5 - 14.4) | 55 (50.9 - 59.2) | 50.2 (45.5 - 54.9) |  | 7.3 (6.6 - 8) | 18.2 (14.7 - 21.7) |  |
| *Sex, %* | | | | | | | |
| Male | 44.5 (43.6 - 45.4) | 33.9 (29.5 - 38.2) | 41.1 (36.8 - 45.4) | ** | 45.8 (44.6 - 47) | 42.8 (37.8 - 47.7) |  |
| Female | 55.5 (54.6 - 56.4) | 66.1 (61.8 - 70.5) | 58.9 (54.6 - 63.2) |  | 54.2 (53 - 55.4) | 57.2 (52.3 - 62.2) |  |
| *Race/ethnicity, %* | | | | | | | |
| Hispanic | 8 (5.8 - 10.2) | 7.2 (3.5 - 10.9) | 6.7 (4.2 - 9.2) |  | 8 (5.9 - 10.1) | 9.6 (5.3 - 14) |  |
| Non-Hispanic Black | 8.8 (7.8 - 9.9) | 13.6 (10.1 - 17) | 12.8 (10 - 15.6) |  | 8.2 (7.1 - 9.3) | 8.9 (6.5 - 11.2) |  |
| Non-Hispanic White | 80.5 (77.9 - 83.2) | 76.7 (71.4 - 82) | 76.3 (72 - 80.6) |  | 81.2 (78.6 - 83.8) | 79.6 (75.2 - 83.9) |  |
| Other | 2.6 (2 - 3.3) | 2.5 (0.7 - 4.4) | 4.2 (1.4 - 7) |  | 2.6 (2 - 3.2) | 1.9 (1.1 - 2.8) |  |
| Greater than high school, % | 35.1 (32.8 - 37.4) | 21.1 (17.6 - 24.7) | 15.4 (11.4 - 19.4) | ** | 37.8 (35.3 - 40.3) | 31.6 (25.3 - 37.8) | * |
| Lives with partner, % | 58.5 (56.8 - 60.2) | 33.5 (28.2 - 38.9) | 35 (29.8 - 40.1) |  | 62.7 (61 - 64.5) | 50.5 (44.4 - 56.6) | *** |
| *Insurance, %* | | | | | | | |
| Dual-eligible | 12.5 (10.9 - 14) | 30.2 (25.4 - 35) | 23.6 (19.5 - 27.7) | ** | 9.6 (8.2 - 11) | 20.6 (16 - 25.2) | *** |
| Medicare Advantage | 41 (38.5 - 43.5) | 40.5 (36.5 - 44.6) | 44.2 (39.2 - 49.2) |  | 40.7 (38.1 - 43.3) | 42.5 (36.9 - 48) |  |
| *Survey response mode, %* | | | | | | | |
| Self respondent | 95.5 (95 - 96.1) | 57.4 (53 - 61.8) | 85.1 (80.4 - 89.8) | *** | 99.2 (99 - 99.4) | 98.8 (97.8 - 99.7) |  |
| Web respondent | 11.2 (10 - 12.5) | 0.6 (-0.3 - 1.6) | 2.9 (0.1 - 5.6) | *** | 12.9 (11.5 - 14.3) | 8.3 (5.1 - 11.5) | ** |
| *Physical health* | | | | | | | |
| Self-reported health, poor or fair | 27.3 (26 - 28.7) | 53.7 (49 - 58.4) | 45.4 (40.1 - 50.7) | ** | 23 (21.7 - 24.3) | 39.4 (34 - 44.9) | *** |
| 1+ ADL limitation | 19.9 (18.7 - 21.1) | 61.9 (57.6 - 66.3) | 41.2 (35.7 - 46.7) | *** | 13.8 (12.5 - 15) | 35.4 (29.9 - 40.9) | *** |
| 1+ IADL limitation | 17.8 (16.8 - 18.8) | 75.6 (71.8 - 79.3) | 51.9 (46.2 - 57.6) | *** | 9.8 (9 - 10.7) | 30.5 (27.4 - 33.7) | *** |
| High blood pressure diagnosis | 66.3 (64.7 - 67.8) | 69.9 (64.8 - 75) | 74.8 (70.6 - 79) |  | 65 (63.2 - 66.7) | 72.1 (67.5 - 76.7) | *** |
| Diabetes diagnosis | 27.7 (26.4 - 29) | 29.6 (26.1 - 33.2) | 36.8 (32.2 - 41.4) | *** | 26.6 (25.3 - 27.9) | 32.1 (26.5 - 37.7) | ** |
| Died during 2018 | 1.7 (1.4 - 2.1) | 10.6 (7.5 - 13.7) | 3.1 (1.3 - 4.9) | *** | 0.7 (0.5 - 1) | 3.9 (2.2 - 5.5) | *** |
| *Cognitive health* | | | | | | | |
| *Self response (N=43,856,771)* | | | | | | | |
| Immediate word recall, mean | 5.4 (5.4 - 5.5) | 2.6 (2.4 - 2.7) | 2.9 (2.8 - 3.1) | *** | 5.8 (5.7 - 5.8) | 5 (4.9 - 5.2) | *** |
| Delayed word recall, mean | 4.5 (4.4 - 4.5) | 1 (0.8 - 1.1) | 1.3 (1.2 - 1.4) | *** | 4.8 (4.8 - 4.9) | 3.9 (3.7 - 4) | *** |
| Serial 7s, mean | 3.7 (3.6 - 3.7) | 1.9 (1.7 - 2.1) | 2.2 (2 - 2.5) | ** | 3.9 (3.8 - 3.9) | 3.3 (3.2 - 3.5) | *** |
| Backwards count, correct 1st attempt, % | 93.4 (92.7 - 94.1) | 75.2 (70.4 - 80) | 82.6 (78.4 - 86.8) |  | 95 (94.2 - 95.7) | 92.6 (90.2 - 95.1) | * |
| Correct date recall (4 of 4), % | 74.4 (72.7 - 76.1) | 16.3 (12 - 20.6) | 37.4 (31.4 - 43.4) | *** | 79.5 (77.6 - 81.4) | 70.3 (65.1 - 75.4) | *** |
| Correct name recall (4 of 4), % | 59.5 (52.8 - 66.2) | 9 (5.6 - 12.4) | 18.4 (14 - 22.7) | *** | 65 (57.7 - 72.2) | 49.2 (42.5 - 55.9) | *** |
| *Proxy response (N=43,856,771)* | | | | | | | |
| IQCODE, mean | 3.8 (3.7 - 3.9) | 4.1 (4 - 4.2) | 3.5 (3.4 - 3.7) | *** | 3 (2.9 - 3.1) | 3.2 (3 - 3.5) | NA |
| Proxy memory score, mean | 4 (3.9 - 4.1) | 4.5 (4.3 - 4.6) | 3.6 (3.3 - 3.9) | *** | 2.5 (2.1 - 2.9) | 3.6 (3.2 - 4.1) | NA |
| Jorm symptom count, mean | 1.5 (1.4 - 1.7) | 2 (1.8 - 2.2) | 0.7 (0.5 - 1) | *** | 0.1 (-0.1 - 0.3) | 0.6 (-0.2 - 1.4) | NA |
| *Setting, %* | | | | | | | |
| Rural | 27.6 (24.3 - 31) | 31.6 (25.4 - 37.9) | 36.1 (28.2 - 44) |  | 27 (23.7 - 30.3) | 24.6 (18.8 - 30.3) |  |
| Urban | 72.4 (69 - 75.7) | 68.4 (62.1 - 74.6) | 63.9 (56 - 71.8) |  | 73 (69.7 - 76.3) | 75.4 (69.7 - 81.2) |  |
| *Census division, %* | | | | | | | |
| New England | 4.5 (1.3 - 7.6) | 4.2 (0.9 - 7.4) | 6.3 (0.3 - 12.3) | * | 4.5 (1.3 - 7.6) | 3 (0.7 - 5.3) |  |
| Middle Atlantic | 11.2 (7.8 - 14.6) | 10.6 (6.1 - 15.2) | 8.1 (5.1 - 11.1) |  | 11.5 (8.1 - 14.9) | 10.8 (6 - 15.6) |  |
| East North Central | 16 (13.5 - 18.5) | 18.8 (13 - 24.7) | 16.2 (11.1 - 21.3) |  | 15.9 (13.6 - 18.2) | 14.3 (9.3 - 19.2) |  |
| West North Central | 8.5 (6 - 11.1) | 7.7 (4.4 - 11.1) | 10 (4.4 - 15.7) |  | 8.5 (6.2 - 10.8) | 9.1 (4.9 - 13.4) |  |
| South Atlantic | 23.4 (20.6 - 26.2) | 22.5 (17 - 28) | 21.4 (15.9 - 27) |  | 23.5 (20.9 - 26.2) | 24.3 (18.5 - 30.1) |  |
| East South Central | 6.4 (4.3 - 8.6) | 8.1 (3.7 - 12.5) | 4.5 (2.3 - 6.8) |  | 6.5 (4.3 - 8.6) | 5.2 (2.6 - 7.9) |  |
| West South Central | 9.8 (7.7 - 11.8) | 13.1 (8.9 - 17.4) | 11 (4.4 - 17.7) |  | 9.2 (7.2 - 11.1) | 12.5 (9 - 16) |  |
| Mountain | 7 (3.8 - 10.3) | 6 (1.2 - 10.8) | 6.8 (1.8 - 11.7) |  | 7.2 (4.1 - 10.4) | 5.8 (2.2 - 9.4) |  |
| Pacific | 13.1 (10.4 - 15.9) | 8.9 (4.3 - 13.5) | 15.5 (9.5 - 21.6) |  | 13.2 (10.8 - 15.7) | 14.9 (9 - 20.8) |  |

Abbreviations: ADL = activities of daily living; CI = confidence interval; GED = general education development; HRS = Health and Retirement Study; IADL = instrumental activities of daily living; IQCODE = Informant questionnaire on cognitive decline in the elderly.

^a^ Reported p-values represent the median of estimates obtained across the 10 imputed datasets, where * = p-value <0.1; ** = p-value <0.05; *** = p-value < 0.01. For proxy-respondent only items (IQCODE, proxy memory score, Jorm symptom count), the TP vs. FP p-values were not estimable because too few survey strata contained information from both groups. Inferences about significance in differences can be made from group means and 95% confidence intervals.

Notes: A subset of self-report cognitive data were missing for wave 2018 web-respondents (11.2% of the sample); we used multiple imputation chained equations (MICE) to generate 10 sets of imputations for these missing data. Point estimates were averaged across the ten datasets, and confidence intervals incorporate both within- and between-imputation variability.

All analyses use complex variance estimation to account for the HRS survey design.

**Appendix 15.** Performance of specific ICD-10-CM Codes Against HRS Dementia

| **ICD code** | **N** | **Percent (95% CI)** | |
| --- | --- | --- | --- |
|  |  | **True Positive** | **False Positive** |
| *HRS LASSO Model Dementia* | | | |
| F03 | 813,186 | 61 (52 - 69) | 39 (31 - 48) |
| F01, F02 and/or G30 | 266,062 | 57 (42 - 71) | 43 (29 - 58) |
| G3184 | 642,429 | 23 (14 - 33) | 77 (67 - 86) |
| G319 | 840,697 | 17 (10 - 24) | 83 (76 - 90) |
| R54 | 255,429 | 39 (22 - 55) | 61 (45 - 78) |
| All other possible category codes | 138,546 | 15 (0 - 29) | 85 (71 - 100) |
| *HRS Modified-Hurd Model Dementia* | | | |
| F03 | 869,198 | 869,198 | 62 (53 - 70) |
| F01F02G30 | 307,798 | 307,798 | 54 (40 - 67) |
| G3184 | 699,972 | 699,972 | 20 (12 - 29) |
| G319 | 881,787 | 881,787 | 20 (11 - 29) |
| R54 | 274,332 | 274,332 | 41 (27 - 55) |
| All other possible codes | 148,239 | 148,239 | 11 (-1 - 24) |
| *HRS Expert Model Dementia, Excluding Web Respondents* | | | |
| F03 | 837,026 | 60 (52 - 69) | 40 (31 - 48) |
| F01F02G30 | 302,764 | 54 (40 - 67) | 46 (33 - 60) |
| G3184 | 612,251 | 27 (18 - 37) | 73 (63 - 82) |
| G319 | 869,393 | 17 (8 - 26) | 83 (74 - 92) |
| R54 | 249,849 | 34 (19 - 48) | 66 (52 - 81) |
| All other possible codes | 125,846 | 16 (1 - 32) | 84 (68 - 99) |

Notes: F01 = Vascular dementia; F02 = Dementia in other diseases classified elsewhere; F03 = Unspecified dementia; G30 = Alzheimer’s disease; G31.84 = Mild cognitive impairment (MCI), so stated; G31.9 = Degenerative disease of nervous system, unspecified; R54 = Age-related physical debility. All other possible category codes include: F04 (Amnestic disorder due to known physiological condition), F06.1 (Catatonic disorder due to known physiological condition), F06.8 (Other specified mental disorders due to known physiological condition), G13.8 (Systemic atrophy primarily affecting central nervous system in other diseases classified elsewhere), G31.2 (Degeneration of nervous system due to alcohol), G31.89 (Other specified degenerative diseases of nervous system), G94 (Other disorders of brain in diseases classified elsewhere).

A subset of self-report cognitive data were missing for wave 2018 web-respondents (11.2% of the sample); we used multiple imputation chained equations (MICE) to generate 10 sets of imputations for these missing data. Point estimates were averaged across the ten datasets, and confidence intervals incorporate both within- and between-imputation variability.

All analyses use complex variance estimation to account for the HRS survey design.
